# Influence of microbial composition and sample type on antimicrobial resistance in urinary tract infections: a single-centre retrospective cohort study (2015–2023)

**DOI:** 10.64898/2026.02.23.26344629

**Authors:** Ashim Kumar Dubey, Jorge Reyes, Clemens Rhiner, Knut Drescher, Jörn Dunkel, John McKinney, Adrian Egli

## Abstract

**Objectives:** To quantify how urine sample type and polymicrobial context impact antimicrobial resistance (AMR) in urinary tract infections (UTIs), using routine diagnostics at scale.

**Methods:** In this retrospective, single-centre study, we analysed 188,687 urine cultures from the Institute of Medical Microbiology, University of Zurich, Switzerland (January 2015 to May 2023). We compared midstream urine (MU), indwelling catheter (IDC), and intermittent catheter (IMC) samples. Samples were classified as negative, bacteriuria, or UTI, by meeting a microbiological UTI threshold (≥10^5^ CFU/mL). We compared sample types using covariate-adjusted regression and constrained ordination for community composition. In bimicrobial cultures, we assessed co-occurrence using adjusted pairwise odds ratios and degree-preserving permutation null models, supported by partner-choice analyses. AMR was modelled as acquired resistance (AR) and total resistance (TR: acquired + intrinsic) probabilities, with predictor contributions quantified using mutual information.

**Results:** Among 186,819 MU, IMC, IDC samples, 56,867 met the UTI threshold. Catheter-associated UTIs (IDC and IMC) were ~60% more likely to be polymicrobial than MU samples. Community composition differed by sample type (p<0·001). In IDC, *Escherichia coli* was less prevalent than in MU, but device-associated pathogens like *Pseudomonas aeruginosa* and *Candida albicans* were enriched. Most species-pairs showed no increased co-occurrence after adjusting for covariates, but a subset showed reproducible enrichment across methods (e.g., *C. albicans-C. glabrata*). Organism identity was the dominant determinant of AMR, with the highest mutual information across AR and TR. AR was higher in IDC for common uropathogens (e.g., *E. coli*). Co-isolation with hospital-associated partners (e.g., *Enterococcus faecium*) was associated with further AR increase. From 2015 to 2023, AR increased from ~48% to ~60%, with rising β-lactam (+β-lactamase inhibitor) resistance and declining fluoroquinolone resistance in *Enterobacterales*.

**Conclusions:** Sample type and co-isolated partners provide clinically actionable information beyond pathogen identity and could support more context-aware reporting and empiric prescribing.

## Introduction

Urinary tract infections (UTIs) are a major healthcare burden, with >400 million infections and ~240,000 deaths estimated in 2019 (1). In hospitalized patients, catheterization amplifies risk: catheter associated UTIs (CAUTIs) account for ~40% of hospital-acquired infections (2) and are difficult to eradicate because of device-associated biofilms and complex microbial ecology (3). CAUTI treatment failures are common, with antibiotic treatment success rates of only ~40% in older adults (4). In addition, multidrug resistance is increasing in uropathogens – UTIs were directly attributable to ~65,000 AMR-attributable deaths in 2019 (5).

Polymicrobial infections are particularly common in catheterized patients (6). In long-term catheterization, up to 80% of bacteriuric episodes are polymicrobial, (7) and polymicrobial bacteraemic UTIs have been associated with increased mortality (8). Beyond the bladder, polymicrobial infections are known to have altered virulence and treatment responses (9–11). In the bladder, catheter surfaces and nutrient-rich urine favour biofilm formation and interspecies interactions that can enhance antibiotic tolerance and persistence (3,11–13). Surveillance data further indicate differences in CAUTIs and non-CAUTIs, both in species composition and resistance profiles (13,14).

Most UTI research is monomicrobial in focus, concentrating on a single pathogen (*Escherichia coli*) and a limited set of resistance phenotypes (15). This approach obscures co-occurrence dynamics of different species, and how partner organisms, sample type, and patient factors jointly shape infection risk and antibiotic susceptibility (6,12). Few studies attempt to integrate ecological models of microbial interactions with patient metadata (age, sex, sample type, etc) at a population-level. Animal data and *in vitro* work show that specific pathogen combinations can display collective antibiotic tolerance and altered virulence (11,13,16,17), but these interactions have rarely been quantified in routine diagnostic data. Most surveillance networks also aggregate catheter and non-catheter urines (14), limiting our ability to distinguish device-specific ecology from host and setting effects. Machine-learning approaches can predict UTI risk from clinical features (18), but do not address the joint influence on resistance.

This study uses a large retrospective dataset from a single centre to address these limitations. We first quantified how catheterization and demographics relate to mono- and polymicrobial infections and overall community composition. We then combined adjusted prevalence ratios, co-occurrence odds ratios, degree-preserving permutations, and directional partner-choice models to characterize organism–organism and organism–sample type associations. Finally, we modelled acquired and total resistance at the species and antibiotic-class level, including partner effects, to identify the main drivers of resistance. Our aim was to provide an ecological view of UTI epidemiology that can inform diagnostics and antimicrobial stewardship.

## Methods

We analysed a routine urine culture and AST database from the Institute of Medical Microbiology (University of Zurich), 2015–2023. We compared sample types using adjusted regression models and constrained ordination, characterised pairwise co-detection patterns using complementary association and null-model approaches, and modelled total vs acquired resistance with covariate adjustment.

Full methodology is in the supplementary materials.

## Results

We analysed data from 188,687 urine cultures (2015–2023), which comprised three different sample types: 132,713 midstream urine (MU) samples, 31,975 indwelling catheter (IDC) samples, and 22,131 intermittent catheter (IMC) samples **(Fig. 1A, B)**. In the absence of symptom data, cultures were classified using standard microbiologic thresholds as negative (no growth, unknown species <10^4^ CFU/mL, samples with no quantification), bacteriuria (<10^5^ CFU/mL), or microbiologically significant UTI (≥10^5^ CFU/mL). Overall, 30·4% of samples met the UTI threshold **(Fig. 1C)**. The proportion of polymicrobial cultures increased with catheterization (IMC 35%, IDC 30%, and MU 18%; **Fig. 1D, Table S1)**. After adjustment for sex, age, ward group, and year, catheter-associated UTI samples were about 60% more likely to be polymicrobial than MU: for IDC the adjusted prevalence ratio (PR) was 1·63 [95% CI 1·56–1·70] (**Table S2)**. Throughout, we distinguish bimicrobial (two species) from those with ≥3 species.

**Figure 1.**
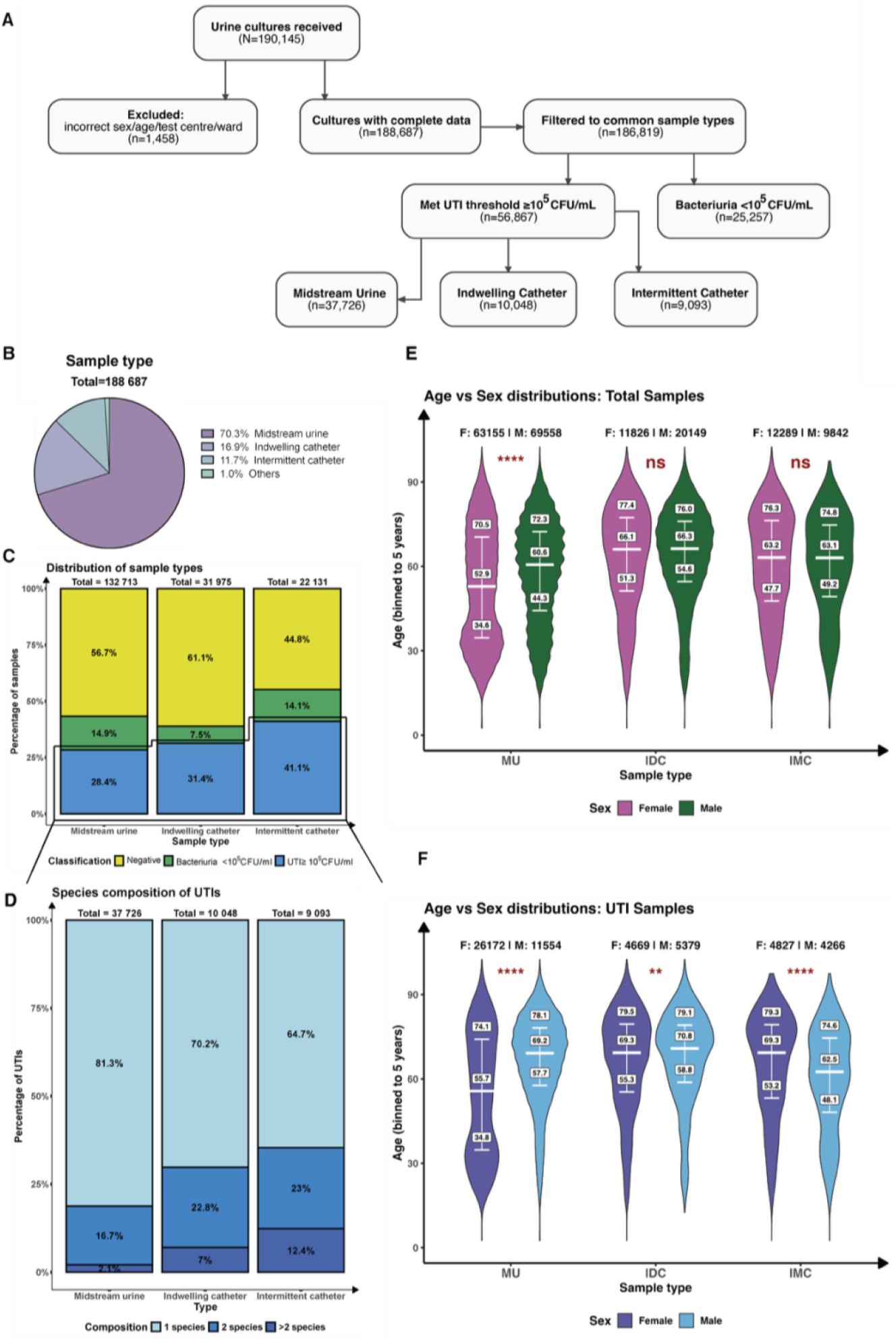
Description of the dataset used in this study. This figure describes the dataset used in this study, beginning with A), showing the sample distribution and classification as a flowchart. Next, we show B) Sample types, describing the proportion of samples from each urine collection type, C) Classification of urine microbial composition data into negative samples, low microbial numbers (in CFU/ml) as bacteriuria and high microbial numbers (in CFU/ml) as UTI, D) Species composition of UTIs into monomicrobial, bimicrobial and polymicrobial UTIs, followed by the binned age-sex distributions of patients for E) Total samples, and F) UTI samples, across sample types. Statistical analysis in E) and F) was performed using a Mann Whitney U-test, where * indicates P < 0·05; **, P < 0·01; ***, P < 0·001 and ****, P < 0·0001.

In tested samples, age distributions were similar by sex for IDC and IMC, though MU showed older males than females (median difference ~8 years; **Fig. 1E**). Among UTI cases, age–sex differences were significant for all three sample types (**Fig. 1F**): MU females were younger than males (in median ~14 years), driven by a large cohort of young female MU patients, which is absent in males. IDC showed a minimal male–female median difference (~1 year), while IMC females were older than males (~7 years). MU specimens predominantly originated from outpatient care, IDC from intensive care, and IMC from external hospitals. For MU and IMC, the distribution of UTI cases mirrored tested samples, whereas UTI IDC specimens, particularly bimicrobial UTIs, were disproportionately referred from external hospitals (including paraplegic units) **(Fig. S1)**. The overall mix of urinary pathogens significantly differed by sample type, even after adjusting for sex, age, ward group, and year of collection (global constrained ordination test p<0·001, sample type explained ~0·7% of compositional variation). Sex and age showed similarly significant associations, ward group a weaker one, and year of collection had no measurable impact. Pairwise analyses indicated that IDC samples differed most from MU samples, whereas IMC samples were compositionally closer to MU than to IDC. Full statistics are shown in **Table S3**.

In monomicrobial infections, *E. coli* was the dominant pathogen across all sample types but was markedly less frequent in IDC than MU (21·9% vs 39·8% (**Fig. 2A**); PR 0·55 [0·52–0·58]; q<0·001) (**Fig. 3A**), with no meaningful enrichment in IMC (43·7%; PR 1·04 [1·00–1·07]. Across sample types, *E. coli* was also consistently less common in males than in females (e.g. MU: 33·4% vs 42·5% (**Figs. S2A**,**B**); PR 0·74 [0·72–0·77]) (**Fig. S3A**), while several non–*E. coli* pathogens were relatively more frequent in male patients (15). For example, *Enterococcus faecalis* showed only mild differences by sample type but was clearly enriched in male patients across MU, IDC, and IMC (e.g. MU: 11·4% in men vs 4·1% in women; PR 2·52 [2·30–2·77]). Classic device-associated organisms were strongly enriched in IDC compared with MU, such as *Candida albicans* (8·6%; PR 5·80 [4·77–7·06]), *Candida glabrata* (3·9%; PR 2·90 [2·34–3·60]), and *Pseudomonas aeruginosa* (6·1%; PR 2·43 [2·10–2·81]). These enrichments were sex-dependent: *P. aeruginosa* was more frequent in male patients (PR 2·07 [1·68-2·54]), whereas both *C. albicans* and *C. glabrata* were more frequent in females. In contrast, presumed commensals such as *Lactobacillus* spp. were strongly depleted in catheterized samples (e.g. IDC: PR 0·18 [0·13-0·25]) and in men (e.g. MU: 0·6% vs 16·6%; PR 0·06 [0·05–0·08]), with *Gardnerella vaginalis* showing similar patterns. *Klebsiella pneumoniae* showed only modest variation, being slightly reduced in IDC but enriched in IMC vs MU. Patterns in bimicrobial infections generally paralleled the monomicrobial findings (**Figs. 3B, S3B**). The most common partner organism was *E. coli* in MU (45·3%) and IMC (48·4%), whereas *E. faecalis* predominated in IDC (38·1%) (**Fig. 2B**), reflecting the underlying sex distribution: *E. coli* was the leading partner in female patients across all sample types, while *E. faecalis* was more frequent as a partner in males in MU and IDC (**Figs. S2C**,**D**).

**Figure 2.**
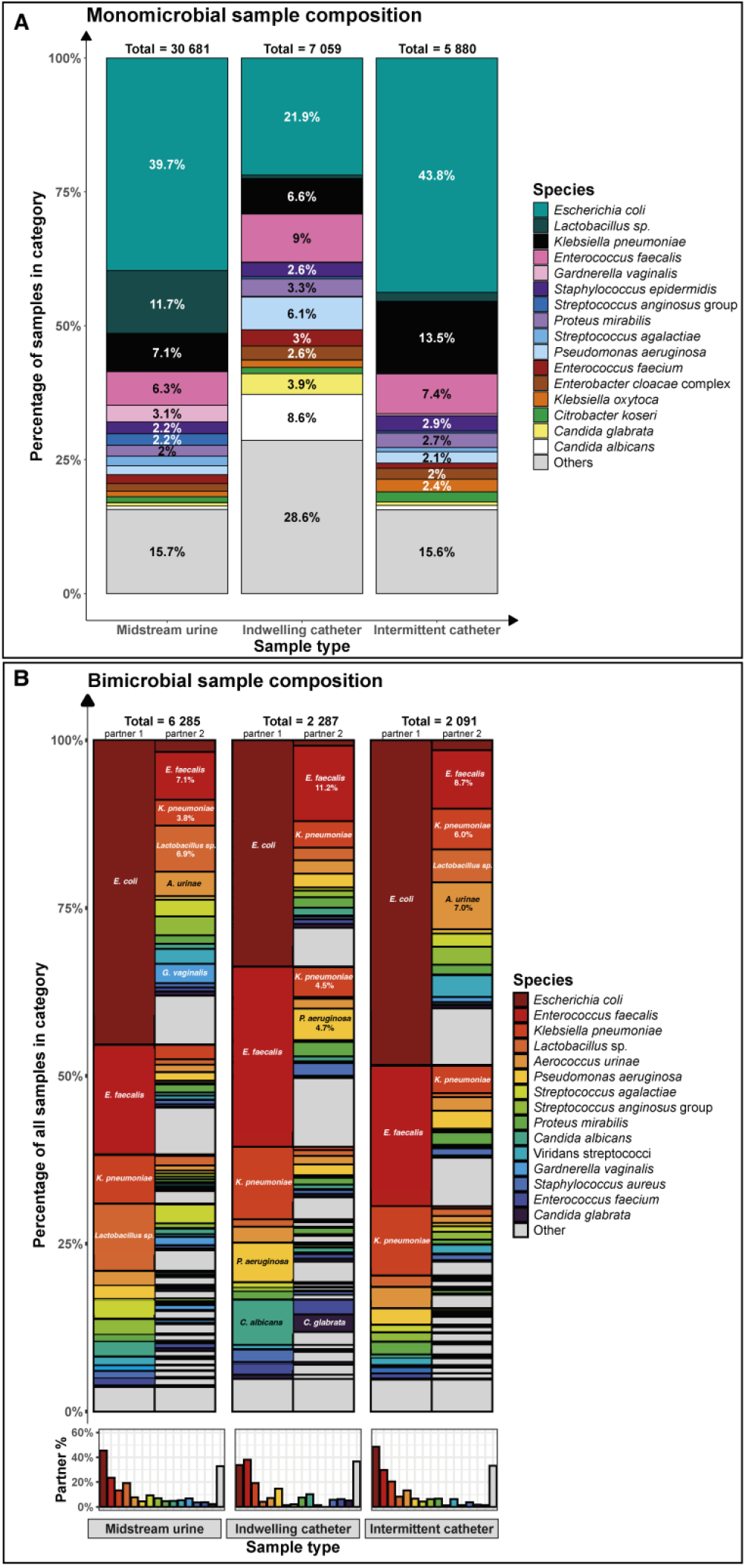
Composition of monomicrobial and bimicrobial UTI samples, based on urine sample type. This figure compares the microbial composition of UTIs, for A) monomicrobial and B) bimicrobial UTI samples. In A), microbes which make up >2% share of the total have their share labelled per sample type. In B), the top 5 microbial pairs are labelled, and the top 3 have their percentage share mentioned. The inset figures at the bottom refer to the percentage of samples, per sample type, that have the organisms graphed as a partner.

**Figure 3.**
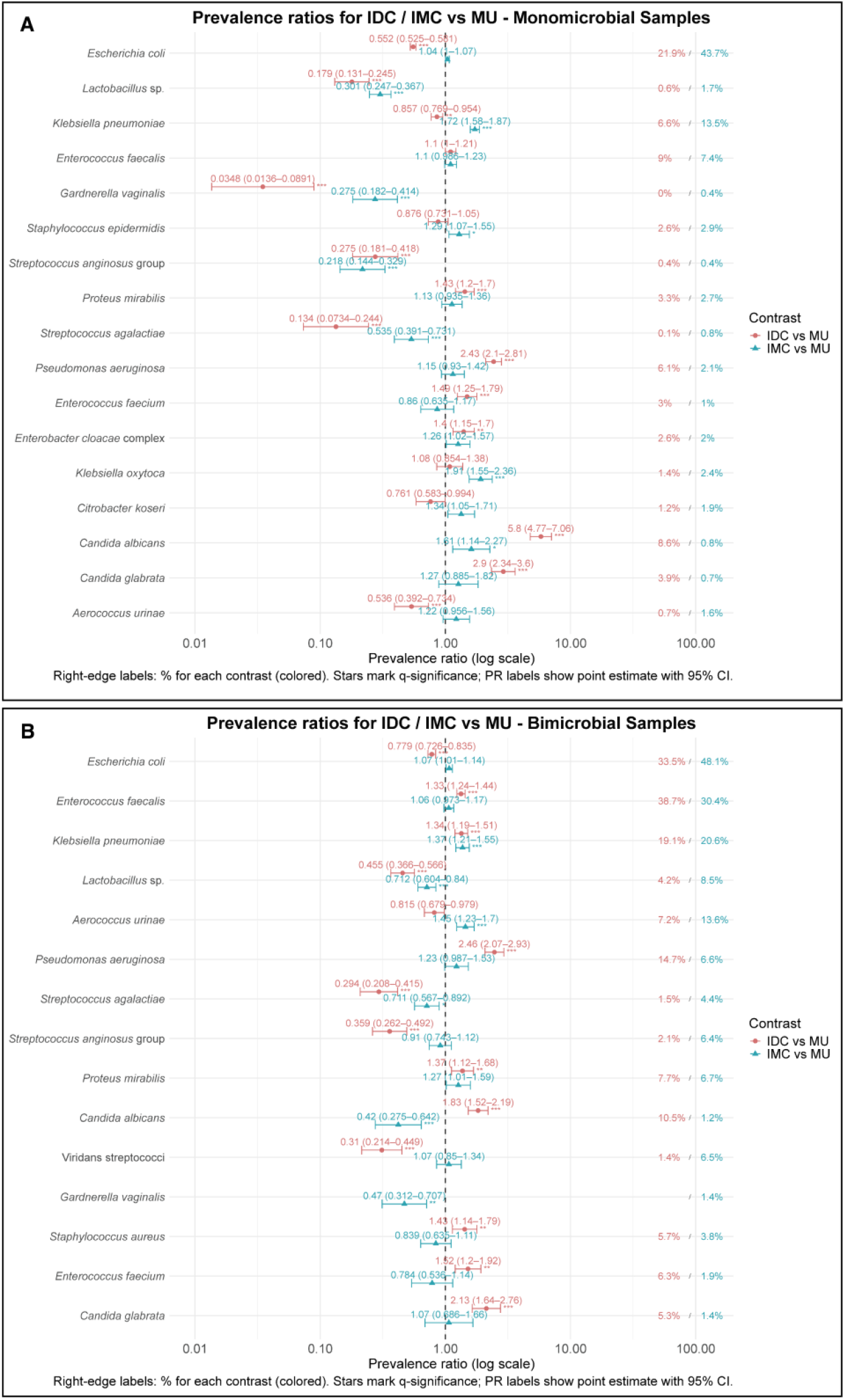
Prevalence ratios (PRs) of species by sample type. Forest plot of PR (point) with 95% CI (horizontal line) for IDC vs MU (in red) and IMC vs MU (in green) on a log scale, with one row per species, for A) Monomicrobial samples, and B) Bimicrobial samples (filtered to 2 unique species per sample). PRs were calculated after adjustment for sex, binned age, ward groups and year of collection. Asterisks denote BH-adjusted significance (*q*<0·05 *, <0·01 **, <0·001 ***). Right-edge labels indicate percent positive in the numerator group (IDC or IMC) to convey base rates. Species plotted are the common species across sample types, like Figure 2. See data files for complete lists.

We next examined which pathogens tend to appear together. Using adjusted co-occurrence odds ratios (ORs) in bimicrobial samples, most organism pairs showed either no clear association or were less likely to co-occur, with only a minority showing increased odds for co-occurrence (**Fig. 4**). These patterns were largely stable in sensitivity analyses that re-fit the model in all UTI samples and further adjusted for species richness (number of species per sample): only four pairs shifted from depletion in bimicrobial samples to enrichment when all samples were included. This discrepancy disappeared once richness was included as a covariate (**Table S4**). This suggests that most signals reflect true pairwise patterns rather than generic polymicrobiality, and these pairs were the exception – they highly co-occur in polymicrobial samples but have negative pairwise associations. Sex-stratified analyses further showed little differences between sexes (**Fig. S4**). Classical co-occurrence tests applied to all samples within each sample type, performed as an independent null check, gave directionally consistent results with the all-sample ORs (**Fig. S5**).

**Figure 4.**
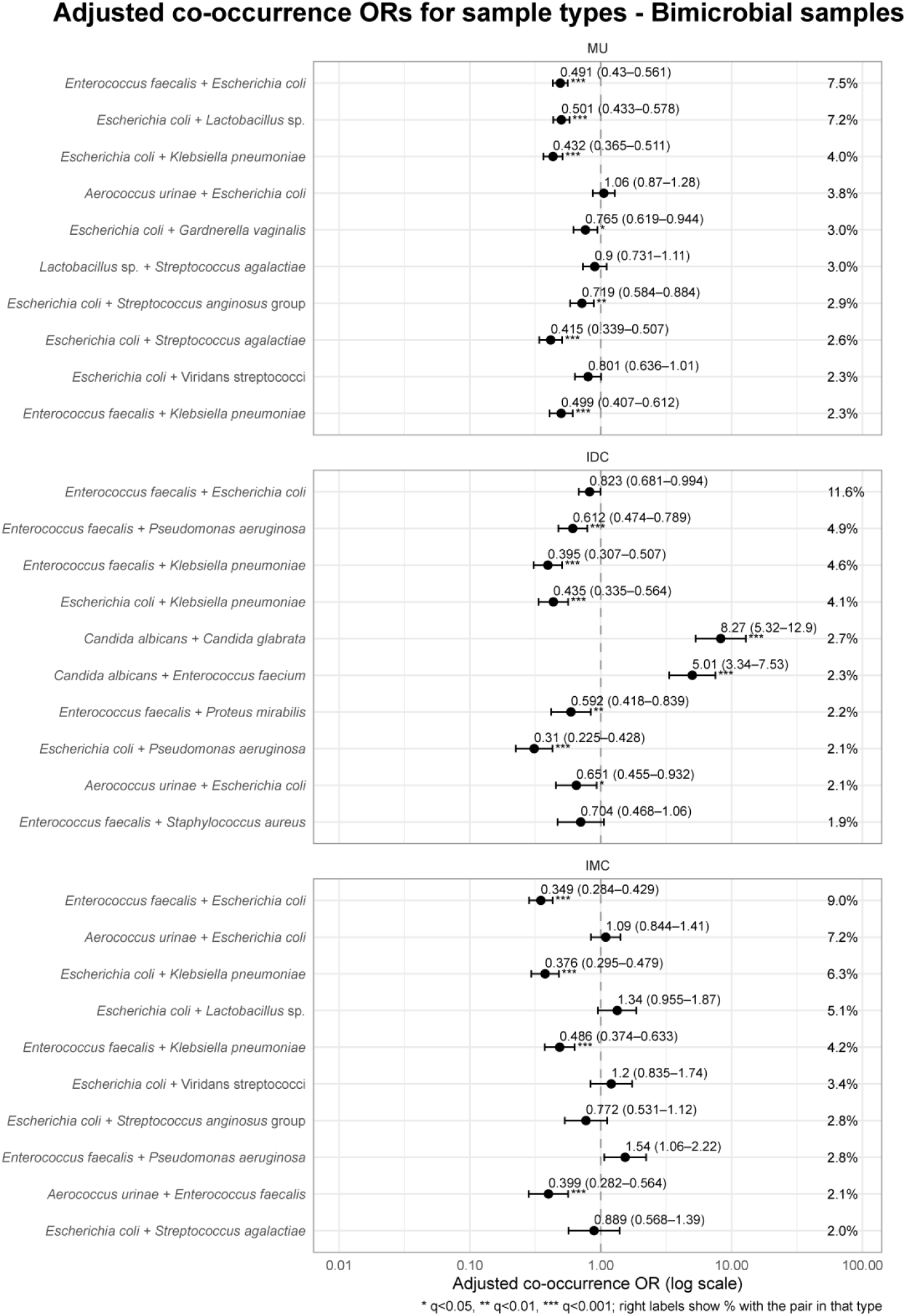
Adjusted co-occurrence odds ratios (OR_sym_) by sample type (Top 10 bimicrobial UTIs per sample type, 2 unique species per sample). Each panel shows OR_sym_ (point) with 95% CI (horizontal line) on a log scale for organism pairs within each sample type. OR_sym_ is the geometric mean of the two directional logistic ORs. CIs use the mean SE on the log scale, and q is the minimum BH-adjusted p across directions. Asterisks mark significance (*q*<0·05 *, <0·01 **, <0·001 ***). Right-edge labels show the pair prevalence (%) (co-appearances / total samples). Models are adjusted for binned age, sex, ward groups and year of collection. See data files for full pair lists.

To understand how background prevalence shapes these species co-occurrence patterns, we performed degree-preserving permutation tests in the bimicrobial subset. Permutation results generally agreed in direction with significant ORs (**Fig. S6, Data files**), with a few discrepancies for significant associations – usually in high prevalence organisms (**Table S5**). For example, the species pair *E. faecalis–E. coli* in MU showed a negative association (OR 0·49), yet the pair still occurred more often than expected under permutation, reflecting their high marginal prevalence. The species pair *E. coli–P. aeruginosa* in MU, however, was strongly depleted by both methods, with far fewer observed pairs than expected and a low OR (0·19), with similar depletions in catheter samples. *C. albicans–C. glabrata* showed a concordant positive signal across methods, with a large OR (OR 8·27), and strong permutation enrichment in IDC, and similar enhancements in MU and IMC. The species pair *C. albicans–Enterococcus faecium* showed a similar pattern of concordant enrichment.

Directional species partner-choice models helped clarify how specific combinations shift across sample types. Among *E. coli* pairs, the proportion that included *E. faecalis* was higher in IDC than MU (risk ratio (RR) 1·68 [1·46–1·94]), whereas among *E. faecalis* pairs the share containing *E. coli* did not increase (RR 0·98 [0·86–1·13]), indicating a specific directionality for the increased IDC prevalence. Similarly, within *E. coli* pairs, the partner mix shifted toward *P. aeruginosa* in IDC vs MU (RR 4·61 [2·78–7·63]), even though absolute co-occurrence of the species pair *E. coli–P. aeruginosa* remained highly depleted by OR. (**Fig. S7**)

We next assessed how sample characteristics relate to the probability that an isolate is antibiotic-resistant, separating acquired resistance (AR) from total resistance (TR, includes intrinsic resistance) (**Fig. 5**). Across sample characteristics, effects were small but potentially clinically relevant. IDC samples had higher TR than MU samples, while AR was broadly similar between the sample types (**Fig. 5A**). We found no evidence of seasonality (**Fig. 5B**). Over the time span 2015–2023, both TR and AR increased, with AR rising from ~48% to ~60%; (**Fig. 5C)**. Sex and age had minor effects. Isolates from male patients had slightly higher TR and slightly lower AR overall, and the oldest age groups showed higher TR with a downward trend in AR (**Fig. 5D–E**). Differences by hospital source were limited, with internal medicine wards showing the highest resistance (**Fig. 5F**). By contrast, organism identity dominated resistance risk. Among the most common pathogens, baseline AR varied widely, from >90% in some species to <10% in others (**Fig. 5G)**. Urine colony count had minimal impact (**Fig. 5H**). Mutual information analysis quantified this contrast: organism identity carried far more information about TR and AR (~0·28 and 0·15 bits, respectively) than any individual patient or specimen variable, each of which contributed <0·01 bits (**Fig. 5I**). Overall, apparent differences by sex, age, or catheterization were largely explained by shifts in the infecting species mix.

**Figure 5.**
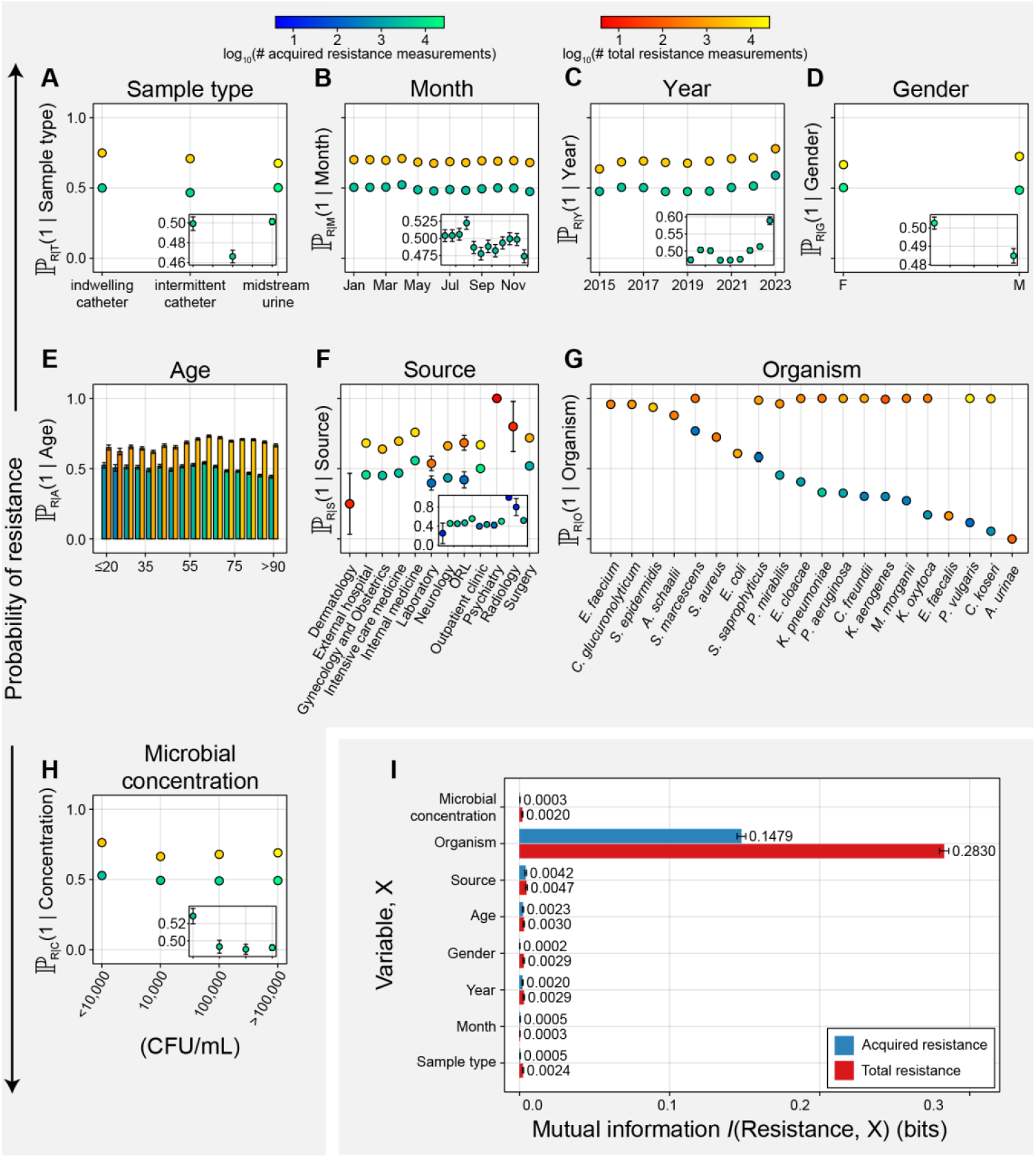
Antibiotic resistance levels vary with sample parameters of patient characteristics, sampling time, and organism. Resistance conditioned on 8 sample metadata is treated as a Bernoulli random variable with parameters as shown in (A)–(H). Samples included in this analysis include all monomicrobial samples which had an antimicrobial susceptibility test (AST) result available. Resistance observed from the ASTs directly were classified as total resistance (red/orange), while resistance data after removing inherent resistances (as defined by EUCAST) were classified as acquired resistances (blue/green). Organisms with no ASTs were not included in this analysis. Sample and patient metadata include (A) sample material, (B) month of sample collection, (C) year of sample collection, (D) patient sex, (E) patient age, (F) medical ward source, (G) organism detected, and (H) concentration of the organism detected. Color scheme legend is shown above plots for the number of samples in each condition. (A)–(H) inset show a magnified view of the acquired resistance plots. (I) **We** measured the dependence between **AMR** and other sample metadata using mutual information which suggests that sample organism is the most predictive variable for antibiotic resistance. All error bars shown are standard error of the mean (SEM). Raw values are included in data files.

Analyses restricted to the most abundant monomicrobial infections were consistent with this picture. IDC samples showed higher AR for common bacteria like *E. coli, K. pneumoniae* and *P. aeruginosa*. Sex differences in AR were small and mainly reflected higher resistance in isolates from male patients for these three species. Age effects varied by organism, but across all strata, between-species variation in AR clearly exceeded the variation attributable to metadata variables (**Fig. S8)**.

Consequently, we then examined AR for individual antibiotics in these species by sample type, calendar month, and year (**Fig. S9**). IDC isolates showed higher resistance across most antibiotic classes, across species. *E. coli* exhibited a striking, systematic increase in resistance to narrow-spectrum β-lactams (ampicillin, amoxicillin– clavulanic acid) over the past few years, a milder decrease in resistance to fluoroquinolones and co-trimoxazole, stable cephalosporin resistance, and rare resistance against carbapenems, nitrofurantoin and fosfomycin. In *K. pneumoniae*, resistance to co-trimoxazole declined, β-lactam/β-lactamase inhibitor resistance increased, and carbapenem resistance stayed low, while cephalosporins and aminoglycosides were stable. In *P. aeruginosa*, class-specific trajectories were relatively stable, but resistance remained consistently higher for fluoroquinolones and carbapenems than in *Enterobacterales*. There was no seasonal variation observed.

Finally, we tested whether resistance changed when isolates were co-detected with specific partners (**Fig. 6**). Most pairings had little effect, but several species-pairings showed partner-associated shifts. *E. coli* was more likely to be resistant when co-isolated with *E. faecium* or *C. albicans*, with increases spanning β-lactam/β-lactamase inhibitor combinations, cephalosporins, fluoroquinolones, and co-trimoxazole, but not fosfomycin or nitrofurantoin (**Fig. 6B, S10**). *K. pneumoniae* tended to be less resistant when paired with *Actinotignum schaalii, Aerococcus urinae*, or *Streptococcus agalactiae*, but more likely to be resistant when paired with P. *aeruginosa* or *E. faecium*, again across multiple drug classes (**Fig. 6C, S10**). For *P. aeruginosa*, co-isolation with *C. albicans* was associated with higher resistance across most antibiotic classes except aminoglycosides (**Fig. 6D, S10**).

**Figure 6.**
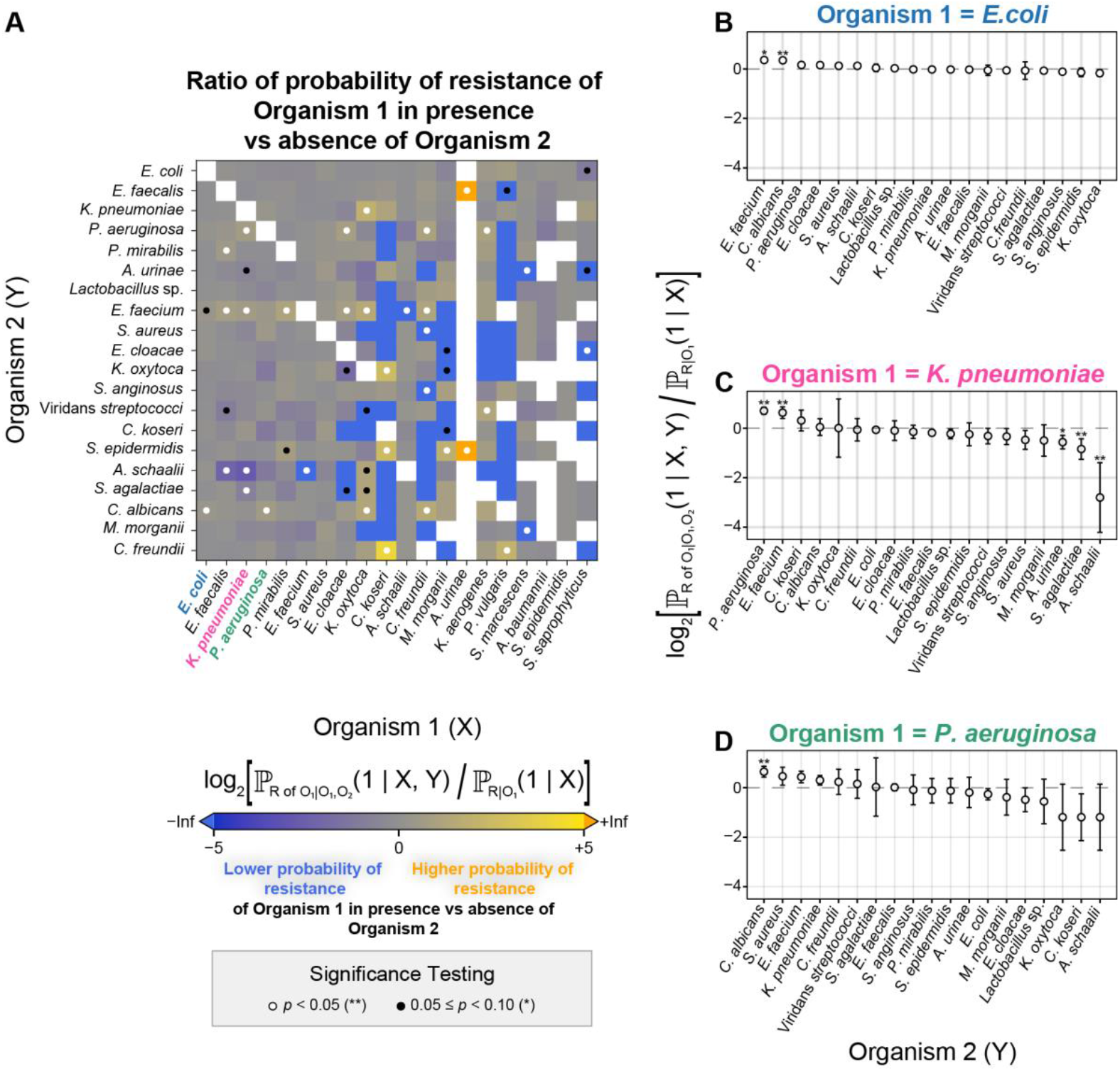
Antibiotic resistance of organisms is influenced by the partner pathogen in bimicrobial samples. The log fold change in resistance of the top 20 most abundant organisms with resistance data in the presence of their partners, which represent the top 20 most abundant organisms in bimicrobial samples are shown as a heatmap (A) with *E. coli* (B), *K. pneumoniae* (C), and *P. aeruginosa* (D) highlighted with partners sorted by descending log-fold change. Error bars are obtained by error propagation of the SEM of the conditioned resistance Bernoulli estimators using the delta method. Significance testing was performed using a double permutation test against a zero-change null hypothesis as described in the Methods section. Raw values are present in data files.

## Discussion

In this single-centre study of urine cultures, we used an ecological framework on routine microbiology data to understand UTI epidemiology and antimicrobial resistance. Catheterization was associated with a higher burden of polymicrobial cultures (6) and enrichment of device-associated pathogens. Co-occurrence analyses showed that most species pairs were neutral or depleted after adjustment, with a small subset of reproducible ecological “hotspots”. The hierarchical analyses help to hypothesize the potential reasons for each pair interaction. Resistance was dominated by organism identity, with sample type and co-isolated partners modulating risk.

While well known for IDC samples, higher polymicrobiality for IMC samples might reflect the use context, and may arise from similar UTI incidence in IDC and IMC (19). Age–sex patterns differed by sample type: younger women contributed disproportionately to MU sample UTIs (20), whereas IMC sample UTIs in comparatively younger male patients likely stem from sex-biased usage, as urinary retention, which increases UTI risk, is >10 times more common in men, with ~60% related to prostatic enlargement (21).

Microbial composition differed clearly between sample types. MU samples were dominated by *E. coli*, though at lower proportions than in community-onset UTI (3), closely resembling healthcare-associated cohorts (13). IDC samples were enriched for device-associated pathogens such as *P. aeruginosa* and *Candida* spp., compatible with catheterization-induced bladder changes, biofilm formation on surfaces and altered antibiotic exposure (22). In contrast, *Lactobacillus* sp. and *G. vaginalis* were largely confined to female MU samples and rare in catheterised urines, consistent with genital tract carry-over during voided collection and bypass during catheterisation. We did not exclude these organisms as “contaminants” due to their morbidity in polymicrobial samples. (23) Although historically labelled as a urinary commensal, (17) *E. faecalis* is now recognized as an important uropathogen, particularly in male patients and CAUTIs (16), and was prominent in our retrospective dataset.

To dissect polymicrobial ecology, we applied a hierarchical framework. PR values captured organism abundances across sample types, OR values quantified pairwise organism-associations, degree-preserving permutations benchmarked pair counts against species abundances, classical *co-occurrence* tests checked independence, and directional partner-choice models assessed shifts in partner mixes relative to MU samples. These complementary approaches showed that catheter context substantially reshapes prevalence and partner selection. Concordant signals (e.g., positive adjusted OR values and permutations) indicated context-specific co-presence beyond marginal frequencies while divergences suggested confounding by shared risk factors. The species pair *E. coli*–*E. faecalis* exemplifies this: it exceeded permutation expectations in MU and IDC samples, but OR values were neutral or negative. Partner-choice explained that the higher prevalence in IDC samples arose from a unilateral shift - while *E. faecalis* occupies a greater share of bimicrobial infections involving *E. coli* than it does in MU, the vice versa isn’t true. This implies overlapping risk profiles (e.g., age, sex, catheter use) and potential asymmetric benefits (nutritional benefits to *E. faecalis* across sample types (7), potential IDC-specific increased tolerance to antibiotics (24) and host immune cells (25) for *E. coli*), rather than symmetric facilitation. The asymmetry might also be limited to be a direct impact of the enhanced PR value in IDC samples for *E. faecalis*. By contrast, the interkingdom species pair *C. albicans-E. faecium* showed concordant enhancement across methods and types, while *C. albicans-E. faecalis* was depleted (26), despite the taxonomic relatedness - underscoring a few ecologically distinct associations warranting mechanistic study.

Resistance was modelled as a Bernoulli outcome and conditioned on sample-level covariates, analysing both TR (clinically actionable for empiric therapy) and AR (relevant to evolution and stewardship). Pathogen identity was the dominant determinant for antibiotic resistance, with patient factors contributing little. Overall, sex and age added limited information, but within the most common uropathogens, we observed higher AR in IDC samples, similar to prior reports (14), and also in male patients, underscoring that where the organism resides adds important information. While there are no seasonal AR variations, unlike prior reports (27), there are large differences in temporal trends for the common uropathogen-antibiotic pairs. Among *Enterobacterales*, AR to amoxicillin-clavulanic acid sharply increased since 2018-19, fluoroquinolone resistance dropped in line with decline in usage and cephalosporin resistance (a proxy for ESBL producing bacteria) plateaued, largely agreeing with the Swiss national trends (28). There were no EUCAST AST guideline changes in this time frame that could explain this, implying a fast-growing resistance, warranting attention.

In polymicrobial infections, the presence of hospital-associated, multidrug resistant or drug-insensitive partners (*E. faecium, P. aeruginosa, C. albicans*) tracked with higher resistance compared to monomicrobial infections, while resistance was unaffected with low-virulence commensals. This pattern, while associative and not causal, suggests that the presence of such organisms may contribute to mark/select resistant lineages, potentially through ecological mechanisms (increased tolerance via drug degradation, biofilm formation, or cross-protection).

This work has a few limitations. First, without symptom or treatment data, some positive cultures likely reflect asymptomatic bacteriuria, despite guidance against sampling without suspected infection, potentially inflating apparent UTI burden (29). Second, our stringent microbiological case definition (≥10^5^ CFU/mL) likely underestimates clinically defined UTI, especially CAUTI (30). As UTI is preceded by colonization, we treated high colonisation as a proxy for UTI, with an emphasis on bimicrobial UTIs. Furthermore, our study is cross-sectional and based on a single centre, so it can identify associations but not temporal sequence or causality, and generalizability beyond this healthcare setting is uncertain. Residual confounding by comorbidity, device dwell time, prior antibiotic exposure, and care pathways is likely, and some low-count strata may remain underpowered despite FDR control.

However, the study also has important strengths: a large, uniformly processed culture dataset, consistent microbiological thresholds, and an explicit ecological framework that links organism identity, sampling context, and community structure, with broad concordance across complementary statistical approaches. Together, our findings suggest that UTI epidemiology and AMR are best understood ecologically: “who” is present, “where” the sample is taken (particularly catheter-associated vs voided urine), and “with whom” the pathogens coexist explain most of the variation in resistance risk, whereas demographic factors contribute comparatively little.

Clinically, this supports interpreting catheter and non-catheter urines differently, treating polymicrobial results as potentially informative rather than mere contamination, and prioritizing pathogen identity and community context when selecting empiric therapy. Future work should validate these patterns prospectively in multicentre cohorts with linked clinical outcomes, dissect key interspecies interactions in mechanistic models, and develop diagnostics including combined ASTs or culture-independent approaches to capture polymicrobial ecology and address the growing challenge of AMR in UTIs.

## Data Availability

All scripts used for analysis in this study are available via GitLab (https://gitlab.epfl.ch/adubey/epi_retro_zurich/), and the output of each statistical analysis is available as accessory data on Zenodo. (https://doi.org/10.5281/zenodo.18338805)

https://doi.org/10.5281/zenodo.18338805

## Contributors

AKD, JM, KD and AE conceptualized the study. JM, KD, AE and JD obtained funding for the study. AKD, JR and CR developed the methodology, did the formal analysis and visualized the data. AKD, CR and AE curated the data. AKD prepared the first draft of the manuscript. All authors revised the content of the manuscript critically and approved the final version. All authors had final responsibility for the decision to submit for publication.

## Data sharing

All scripts used for analysis in this study are available via GitLab (https://gitlab.epfl.ch/adubey/epi_retro_zurich/) and associated data is available on Zenodo. (https://doi.org/10.5281/zenodo.18338805)

## Acknowledgments

This project was supported as part of NCCR AntiResist, a National Center of Competence in Research, funded by the Swiss National Science Foundation (grant number 180541), and an endowment to Prof. Adrian Egli from University of Zurich. We acknowledge the usage of ChatGPT 5.1 for language and coding support and take full responsibility for the accuracy of the work. We also thank past and current members of the Egli, McKinney, Drescher, and Dunkel groups, for their help and guidance for this work.

## References

1. Zeng Z, Zhan J, Zhang K, Chen H, Cheng S. Global, regional, and national burden of urinary tract infections from 1990 to 2019: an analysis of the global burden of disease study 2019. World J Urol. 2022 Mar 1;40(3):755–63.

2. Lo E, Nicolle LE, Coffin SE, Gould C, Maragakis LL, Meddings J, et al. Strategies to Prevent Catheter-Associated Urinary Tract Infections in Acute Care Hospitals: 2014 Update. Infect Control Hosp Epidemiol. 2016/05/10 ed. 2014;35(5):464–79.

3. Flores-Mireles AL, Walker JN, Caparon M, Hultgren SJ. Urinary tract infections: epidemiology, mechanisms of infection and treatment options. Nat Rev Microbiol. 2015 May 1;13(5):269–84.

4. Langford BJ, Daneman N, Diong C, Lee SM, Fridman DJ, Johnstone J, et al. Antibiotic Selection and Duration for Catheter-Associated Urinary Tract Infection in Non-Hospitalized Older Adults: A Population-Based Cohort Study. Antimicrob Steward Healthc Epidemiol. 2023 Jan;3(1):e132.

5. Murray CJL, Ikuta KS, Sharara F, Swetschinski L, Aguilar GR, Gray A, et al. Global burden of bacterial antimicrobial resistance in 2019: a systematic analysis. The Lancet. 2022 Feb 12;399(10325):629–55.

6. Gaston Jordan R., Johnson Alexandra O., Bair Kirsten L., White Ashley N., Armbruster Chelsie E. Polymicrobial Interactions in the Urinary Tract: Is the Enemy of My Enemy My Friend? Infect Immun. 2021 Mar 17;89(4):10.1128/iai.00652-20.

7. Nye TM, Zou Z, Obernuefemann CLP, Pinkner JS, Lowry E, Kleinschmidt K, et al. Microbial co-occurrences on catheters from long-term catheterized patients. Nat Commun. 2024 Jan 2;15(1):61.

8. Siegman-Igra Y, Kulka T, Schwartz D, Konforti N. Polymicrobial and monomicrobial bacteraemic urinary tract infection. J Hosp Infect. 1994 Sept 1;28(1):49–56.

9. Brogden KA, Guthmiller JM, Taylor CE. Human polymicrobial infections. The Lancet. 2005 Jan 15;365(9455):253–5.

10. Nabb DL, Song S, Kluthe KE, Daubert TA, Luedtke BE, Nuxoll AS. Polymicrobial Interactions Induce Multidrug Tolerance in Staphylococcus aureus Through Energy Depletion. Front Microbiol [Internet]. 2019 Dec 5 [cited 2025 Dec 22];10. Available from: https://www.frontiersin.org/journals/microbiology/articles/10.3389/fmicb.2019.02803/full

11. Peters BM, Jabra-Rizk MA, O’May GA, Costerton JW, Shirtliff ME. Polymicrobial Interactions: Impact on Pathogenesis and Human Disease. Clin Microbiol Rev [Internet]. 2012 Jan [cited 2025 Dec 22]; Available from: https://journals.asm.org/doi/10.1128/cmr.00013-11

12. de Vos MGJ, Zagorski M, McNally A, Bollenbach T. Interaction networks, ecological stability, and collective antibiotic tolerance in polymicrobial infections. Proc Natl Acad Sci. 2017 Oct 3;114(40):10666–71.

13. Tandoğdu Z, Bartoletti R, Cai T, Çek M, Grabe M, Kulchavenya E, et al. Antimicrobial resistance in urosepsis: outcomes from the multinational, multicenter global prevalence of infections in urology (GPIU) study 2003–2013. World J Urol. 2016 Aug 1;34(8):1193–200.

14. D’Incau S, Atkinson A, Leitner L, Kronenberg A, Kessler TM, Marschall J. Bacterial species and antimicrobial resistance differ between catheter and non–catheter-associated urinary tract infections: Data from a national surveillance network. Antimicrob Steward Healthc Epidemiol. 2023 Jan;3(1):e55.

15. Heijer CDJ den, Penders J, Donker GA, Bruggeman CA, Stobberingh EE. The Importance of Gender-Stratified Antibiotic Resistance Surveillance of Unselected Uropathogens: A Dutch Nationwide Extramural Surveillance Study. PLOS ONE. 2013 Mar 29;8(3):e60497.

16. Codelia-Anjum A, Lerner LB, Elterman D, Zorn KC, Bhojani N, Chughtai B. Enterococcal Urinary Tract Infections: A Review of the Pathogenicity, Epidemiology, and Treatment. Antibiot Basel Switz. 2023 Apr 19;12(4).

17. Kline KA, Lewis AL. Gram-Positive Uropathogens, Polymicrobial Urinary Tract Infection, and the Emerging Microbiota of the Urinary Tract. In: Urinary Tract Infections [Internet]. 2017 [cited 2023 Sept 5]. p. 459–502. Available from: 10.1128/9781555817404.ch19

18. Møller JK, Sørensen M, Hardahl C. Prediction of risk of acquiring urinary tract infection during hospital stay based on machine-learning: A retrospective cohort study. PLOS ONE. 2021 Mar 31;16(3):e0248636.

19. Neumeier V, Stangl FP, Borer J, Anderson CE, Birkhäuser V, Chemych O, et al. Indwelling catheter vs intermittent catheterization: is there a difference in UTI susceptibility? BMC Infect Dis. 2023 Aug 2;23:507.

20. Hooton TM, Scholes D, Hughes JP, Winter C, Roberts PL, Stapleton AE, et al. A prospective study of risk factors for symptomatic urinary tract infection in young women. N Engl J Med. 1996 Aug 15;335(7):468–74.

21. Yenli EMT, Aboah K, Gyasi-Sarpong CK, Azorliade R, Arhin AA. Acute and chronic urine retention among adults at the urology section of the Accident and Emergency Unit of Komfo Anokye Teaching Hospital, Kumasi, Ghana. Afr J Urol. 2015 June 1;21(2):129–36.

22. Delnay KM, Stonehill WH, Goldman H, Jukkola AF, Dmochowski RR. Bladder histological changes associated with chronic indwelling urinary catheter. J Urol. 1999 Apr 161(4):1106–8 discussion 1108-1109.

23. Gilbert NM, O’Brien VP, Lewis AL. Transient microbiota exposures activate dormant Escherichia coli infection in the bladder and drive severe outcomes of recurrent disease. PLOS Pathog. 2017 Mar 30;13(3):e1006238.

24. McMurtry TA, Barekat A, Rodriguez F, Purewal P, Bulman ZP, Lenhard JR. Capability of Enterococcus faecalis to shield Gram-negative pathogens from aminoglycoside exposure. J Antimicrob Chemother. 2021 July 10;76(10):2610–4.

25. Tien BYQ, Goh HMS, Chong KKL, Bhaduri-Tagore S, Holec S, Dress R, et al. Enterococcus faecalis Promotes Innate Immune Suppression and Polymicrobial Catheter-Associated Urinary Tract Infection. Infect Immun. 2017 Nov 17;85(12):10.1128/iai.00378-17.

26. Cruz MR, Graham CE, Gagliano BC, Lorenz MC, Garsin DA. Enterococcus faecalis Inhibits Hyphal Morphogenesis and Virulence of Candida albicans. Infect Immun. 2013 Dec 20;81(1):189–200.

27. Bock L, Aguilar-Bultet L, Egli A, Battegay M, Kronenberg A, Vogt R, et al. Air temperature and incidence of extended-spectrum beta-lactamase (ESBL)-producing Enterobacteriaceae. Environ Res. 2022 Dec 1;215:114146.

28. Federal Office of Public Health (FOPH), Federal Food Safety and Veterinary Office (FSVO). Swiss Antibiotic Resistance Report 2024: Usage of Antibiotics and Occurrence of Antibiotic Resistance in Switzerland [Internet]. Bern: Federal Office of Public Health (FOPH); Federal Food Safety and Veterinary Office (FSVO); 2024 Nov [cited 2025 Sept 25]. Available from: https://www.anresis.ch/wp-content/uploads/2024/11/SARR24.pdf

29. Kass EH. Asymptomatic infections of the urinary tract. Trans Assoc Am Physicians. 1956;69:56–64.

30. Hernández-Hernández D, Padilla-Fernández B, Ortega-González MY, Castro-Díaz DM. Recurrent Urinary Tract Infections and Asymptomatic Bacteriuria in Adults. Curr Bladder Dysfunct Rep. 2022;17(1):1–12.

