## Supplementary Files for "Influence of microbial composition and sample type on antimicrobial resistance in urinary tract infections: a single-centre retrospective cohort study (2015–2023)"

### Table of Contents


| Content | Page no. |
|---|---|
| **Methods:** | 3-9 |
| 1 Dataset of routine urine cultures in Zurich (2015-2023) | 3 |
| 2 Statistical analyses | 3-6 |
| 2.1 Polymicrobial Prevalence Ratio analysis | 3 |
| 2.2 Global Composition Analysis | 4 |
| 2.3 Species-specific Prevalence Ratio analysis: | 4 |
| 2.3.1 By sample type | 4 |
| 2.3.2 Male vs female within sample type | 4 |
| 2.4 Co-occurrence Odds-Ratio analysis | 4 |
| 2.5 Classical species cooccurrence (hypergeometric null) | 5 |
| 2.6 Degree-preserving permutation (pairing-null) analysis | 5 |
| 2.7 Partner choice analysis | 6 |
| 3 Resistance Analysis | 7-9 |
| 3.1 Treating Measurements of Antibiotic Resistance as Bernoulli Random Variables | 7 |
| 3.2 $\chi$2 Permutation Analysis for Statistical Significance of Resistance and Susceptibility | 8 |
| 3.3 Calculating Mutual Information | 8 |
| 3.4 Error Propagation via Delta Method for Log-Fold Change | 9 |
| 3.5 $\chi$2 Permutation Analysis for Statistical Significance of Bernoulli Estimator Differences | 9 |
| **Supplementary Figures:** | 10-19 |
| Figure S1 – Origin of urine samples, separated by sample type and category of samples. | 10 |
| Figure S2 – Composition of monomicrobial and bimicrobial UTI samples, based on urine sample type, across male and female patients | 11 |
| Figure S3 – Male vs Female PRs within sample types | 12 |
| Figure S4 – Adjusted co-occurrence odds ratios (ORsym) by sample type (bimicrobial UTIs, stratified by sex) | 13 |
| Figure S5 – Classical co-occurrences in UTIs from midstream urine samples agree with all-sample OR values | 14 |
| Figure S6 – Degree-preserving permutation results in bimicrobial samples. | 15 |
| Figure S7 – Partner choice analysis for E. coli and E. faecalis as partners in bi-microbial UTIs (binary “B vs not-B”) | 16 |
| Figure S8 – Antibiotic resistance prevalence for organisms varies across sample parameters of patient characteristics, sample type and sampling time | 17 |
| Figure S9 – Antibiotic resistance against commonly used antibiotics changes over time, across sample types and for different organisms | 18 |
| Figure S10 – Antibiotic resistance changes in the presence of partners compared to their absence for pathogen combinations with significant differences | 19 |
| **Supplementary Tables:** | 20-21 |
| Table S1: Sex of the sample provider, and number of species in each sample, separated by sample type | 20 |
| Table S2: Prevalence Ratios for polymicrobial nature of samples – adjusted for age, sex, ward and year of collection | 20 |
| Table S3: RDA analysis data – marginal and pairwise, showing the differences between the sample types in terms of microbial composition. | 20 |
| Table S4: This table shows the ORs of species pairs which are significant but non-concordant between the bimicrobial cohort and ORs from the entire UTI database | 21 |
| Table S5: This table shows the species pairs for which the degree preserving permutations are significant but non-concordant to significant adjusted ORs | 21 |
| **References** | 22 |

### Methods

#### 1 Dataset of routine urine cultures in Zurich (2015-2023)

We analysed a deduplicated retrospective dataset from the Institute of Medical Microbiology, University of Zurich (IMM, UZH), containing sample ID, date of collection, patient sex, age, microbiological results (organisms and semi-quantitative counts), and antimicrobial susceptibility test (AST) results of each pathogen.

Data were cleaned to harmonise organism names, antibiotic names, and other metadata. Entries with impossible values or missing key fields (e.g. invalid age, unclassified sex, test cases) were removed. Ward information and identifiers for patients and samples were anonymised. The final analytic dataset comprised 188,687 samples.

The Kantonale Ethikkommission Zürich (KEK Zürich) reviewed the project (BASEC-ID Req-2026-00163) and issued a clarification of responsibility stating that the study does not fall under the Swiss Human Research Act and that ethical approval is not required because it uses anonymised, already existing health-related data.

The analysis of the database was performed primarily in R v4·3·2 (1), with data wrangling and plotting via the tidyverse (2) figure assembly using ggrepel (3) and cowplot (4) packages. Colour palettes were picked from viridis (5), Rcolourbrewer (6) and from Martin Krzwynski's 15 colour palette for colour blindness (7).

#### 2 Statistical analyses

Unless stated otherwise, the parameter "*sample type*" comprised three classes: midstream urine (MU), indwelling catheter (IDC), and intermittent catheter (IMC). The parameter "*sex*" was encoded as Female/Male. The parameter "*age*" was converted to the midpoint of 5-year bins. The parameter "*ward*" was coarsened into six groups (Outpatient, Internal Medicine, ICU, External Hospital, Gynaecology/Obstetrics, Other) to stabilize estimation, and the calendar "*calendar year*" was included as a median-centred numeric covariate (year_c).

We first quantified the sample type differences through polymicrobial adjusted prevalence ratios and constrained ordination-based global composition analysis. We then applied a hierarchical framework to dissect polymicrobial ecology: PR values captured abundances across sample types, adjusted co-occurrence odds ratios (ORs) quantified pairwise-associations, degree-preserving permutations benchmarked pair counts against species abundances, classical co-occurrence tests checked independence, and directional partner-choice models assessed shifts in partner mixes relative to MU samples through risk ratios (RRs).

##### 2.1 Polymicrobial Prevalence Ratio analysis:

We assessed whether the *sample type* is associated with the probability that a sample is polymicrobial after adjusting for covariates. Analyses used the positive samples cohort (to control for sample load), filtered to have only unique species per sample. For each sample we counted the number of unique species and defined a binary outcome:

$$\mathrm{Poly} = 1\left\{n_{unique_{species}} \geq 2\right\} \in \{0,1\},$$

So, polymicrobial means >= 2 species, and monomicrobial means 1 species.

Samples missing any model covariate (*sample type*, *sex*, *age*, *ward*, *year*) were excluded.

Prevalence ratios quantify the prevalence of an outcome (here, sample being polymicrobial) in an exposure group (here, sample types of IDC or IMC), as compared to its prevalence in a reference group (here, MU).

We estimated adjusted prevalence ratios (PRs) for IDC vs MU and IMC vs MU using Poisson regression with a log link and heteroskedasticity-robust (HC0) standard errors (SEs):

$$\log\{\Pr(\mathrm{Poly} = 1)\} = \beta_0 + \beta_{\mathrm{IDC}}\,1(\mathrm{IDC}) + \beta_{\mathrm{IMC}}\,1(\mathrm{IMC}) + \beta_{\mathrm{sex}}\mathrm{sex} + \beta_{\mathrm{age}}\,\mathrm{age} + \beta_{\mathrm{year}}\mathrm{year_c} + \sum_{g} \beta_{\mathrm{ward},g} 1(ward_{group} = g)$$

Exponentiated sample-type coefficients give adjusted PRs and 95% CIs for IDC vs MU and IMC vs MU. P-values for the two sample-type contrasts were Benjamini–Hochberg (BH) adjusted within analysis. All computations were performed using sandwich (8) and lmtest (9) packages in R.

**2.2 Global Composition Analysis:**

To test whether overall urinary community composition differed by sample parameters, we used redundancy analysis (RDA) on presence/absence matrices. Analyses were performed in the UTI cohort after removing samples with multiple morphotypes so every polymicrobial sample had unique species. For each analysis subset, we constructed a sample x species 0/1 matrix, excluded ultra-rare species (prevalence <0·5%), and removed all-zero samples. Samples missing *sample type, sex, age, ward*, or *year* were excluded.

Matrices were Hellinger-transformed (square-root of relative abundances on the 0/1 data), then analyzed using RDA with formula:

$$Y_{Hellinger} \sim sample\ type + sex + age + ward_{group} + year_c$$

Significance was evaluated by permutation ANOVA (999 permutations; marginal tests for each term). We report model adjusted $R^2$ for the full model and *partial* adjusted $R^2$ for each term from partial RDA by conditioning on the other terms (e.g., Y ~ *sample type* + Condition(*sex*) + Condition(*age*) + Condition(*ward_group*) + Condition(*year_c*) for the sample-type partial).

For pairwise comparisons (MU vs IDC, MU vs IMC, IDC vs IMC), we repeated the same preprocessing and model and reported F, *p* (999 permutations; BH-adjusted across the three tests), and adjusted $R^2$ as an interpretable effect size. P-values across the three pairwise tests were BH-adjusted. This analysis was performed using the vegan package (10) in R.

**2.3 Species-specific Prevalence Ratio analysis:**

**2.3.1 By sample type**

For each sample, each species was coded as present/absent (one binary outcome per species). Analyses were run separately for monomicrobial and bimicrobial UTI cohorts. For each species, we estimated PRs for IDC vs MU and IMC vs MU using Poisson regression with log link and HC0 robust SEs:

$$\log\{\Pr(Y_{species} = 1)\} = \beta_0 + \beta_{\text{IDC}}\,1(\text{IDC}) + \beta_{\text{IMC}}\,1(\text{IMC}) + \beta_{\text{sex}}\text{sex} + \beta_{\text{age}}\,\text{age} + \beta_{\text{year}}\text{year}_{\text{c}} + \sum_{g} \beta_{\text{ward},g} 1(ward_{group} = g)$$

For sparse species that passed our filters, we allowed the *ward* and then *year* to be dropped if they caused non-convergence, but no such cases were observed. We used bias-reduced Poisson (brglm2, mean-bias reduction) as the first engine, with standard Generalized Linear Model (GLM) with robust SEs used in case the former fails. We only estimated contrasts when overall species prevalence exceeded ≥0·1%, and there were at least three positives in each contrasted sample type. Contrasts failing thresholds were excluded.

**2.3.2 Male vs female within sample type**

Within each "*sample type*" (MU/IDC/IMC), we compared prevalence in Male vs Female for each species,

$$\log\{Pr\ (Y_{species} = 1)\} = \alpha_0 + \alpha_{\text{Male}}\,1(\text{Male}) + \alpha_{\text{age}}\,\text{age} + \alpha_{\text{year}}\text{year}_{\text{c}} + \sum_{g} \alpha_{\text{ward},g} 1(ward_{group} = g)$$

We required N≥50 in the *sample type* and ≥3 positives per *sex*. Two-sided *p*-values were BH-adjusted within each contrast (*q*), and significance was defined at *q*<0·05 as above. The analysis was performed using the sandwich, (8) lmtest (9), the brglm2 (11) packages in R.

**2.4 Co-occurrence Odds-Ratio analysis:**

For each organism pair $(A, B)$, we estimated the conditional association between their presences in the same sample: the odds ratio for $B$ given $A$, adjusted for covariates and evaluated within strata (overall; by *sample type*;

and by *sample type* × *sex*). The analysis used the sample-level presence/absence matrix (one binary column per species) constructed previously. Within each stratum, we modelled B as the binary outcome and A as the exposure using logistic regression with covariate adjustment:

$$\text{logit}\{Pr\ (B = 1)\} = \alpha + \beta \text{A} + \gamma^{\top}\text{Z}$$

where $Z$ included binned *age*, *sex*, *year_c* and *ward group* (*sex* was excluded in *sex*-stratified models). To capture directionality, we fit both B|A and A|B and then summarized each pair with a symmetric OR,

$$\log(OR_{sym}) = \frac{1}{2}\{\log(OR_{A\to B}) + \log(OR_{B\to A})\}$$

Before modelling a pair in each stratum, we required: species to have overall prevalence > 1%, at least 50 samples per stratum, at least 5 positives for each species in the stratum and at least 5 samples with both present. Pairs failing any screen were recorded as skipped and not analysed.

Models used HC0 robust SEs for Wald inference. For complete or near separation (2×2 tables with zero cells), we attempted mean-bias-reduced logistic regression via brglm2 (Firth-like), falling back to standard GLM if needed. 95% CIs were approximated from the mean SE on the log scale. For multiplicity, we used the minimum BH-adjusted $p$-value across the two directions ($q_{min}$, or q), with the significance threshold set at q< 0·05·

Our primary co-occurrence analysis focused on sample-type strata in strictly bimicrobial UTIs (exactly two unique organisms per sample). As sensitivity analyses, we repeated the same procedure in the full UTI dataset (all richness levels, adjusted for the same covariates), fitted models additionally adjusting for richness ($n_{species}$), and stratified by *sample type* × *sex*. The analysis was performed using sandwich, (8) lmtest (9), the brglm2 (11) packages in R.

**2.5 Classical species cooccurrence (hypergeometric null):**

To test co-occurrence against a simple independence null based on marginal prevalences, we performed the classical cooccur analysis separately within each sample type. For each sample type, we constructed an organism × samples 0/1 presence/absence matrix and fitted cooccur (type = "spp_site", thresh = TRUE, spp_names = TRUE, eff_standard = TRUE, eff_matrix = TRUE). For each organism pair $(i, j)$, the null hypothesis is species–sample independence, with expected co-occurrence

$$E_{ij} = \frac{n_i n_j}{N}$$

Where $n_i$ and $n_j$ are the numbers of samples positive for each organism and N is the number of samples in that *sample type*. Exact tail probabilities are computed from the hypergeometric distribution and reported as $p_{gt}$ (more than expected) and $p_{lt}$ (fewer than expected). We report the species–species co-occurrence heatmap for midstream urine and the observed-vs-expected scatter with the 1:1 line indicating the null expectation – raw data for all sample types is present in the data files. This analysis was performed using the cooccur package (12) in R.

**2.6 Degree-preserving permutation (pairing-null) analysis**:

To separate true pairwise affinity from effects of marginal prevalence and fixed "two-per-sample" structure, we performed a degree-preserving permutation analysis in bimicrobial samples (exactly two distinct organisms per sample). Analyses were stratified by *sample type* and by *sample type* × *sex*.

Each bimicrobial sample contributes one unordered pair (A|B). For each stratum, we computed observed pair counts $C_{AB}$, the degree of each organism $c_i$ (number of times organism $i$ appears across all pairs), and the number of pairs $M$. We then generated a null distribution by random partner-swapping (2-switch/edge-swap Markov chain Monte Carlo) that preserves the degree vector $\{c_i\}$ and the two-per-sample structure, while forbidding self-pairs. Strata with <10 pairs were skipped.

We used a burn-in of BURN_IN_FACTOR × M swaps, then, for each of N_PERMS = 10,000 iterations, performed STEPS_PER_PERM_FACTOR × M successful swaps, recomputed pair counts, and accumulated summary statistics. For each pair we estimated the permutation mean $\mu_{perm}$, SD $\sigma_{perm}$, and reported a Z-score:

$$Z_{perm} = \frac{C_{AB} - \mu_{perm}}{\sigma_{perm}}$$

Two-sided empirical *p*-values used tail counts with a +1 correction, and BH FDR was applied within each stratum ($\alpha = 0{\cdot}05$).

We also report an unconditional expected value,

$$\text{Expected}_{\text{uncond}} = \frac{n_a n_b}{2N - 1}$$

and a shrunk log2 ratio,

$$Value = \log_2 \frac{(\text{Obs} + 1)}{(\text{Expected}_{\text{uncond}} + 1)}$$

As the permutation null conditions on organism prevalence and the pair structure does not adjust for *age* or *sex*, we used it to check concordance with the co-occurrence ORs and to identify pairs whose results could be explained by prevalence. The analysis used data.table (13) and future/future.apply (14) packages in R.

**2.7 Partner choice analysis**:

To assess how *sample type* alters the partner mix of a focal organism A, we modelled the parameter "*partner choice*" in bimicrobial samples. For each bivariable sample we formed an unordered pair and retained it if it contained organism A (*Escherichia coli* or *Enterococcus faecalis*). The other organism was defined as the partner B.

Within each contrast (IDC vs MU or IMC vs MU), we restricted to samples in the respective comparison and selected the top five partners by prevalence in the catheter group. For a partner B, we defined the binary outcome.

$$Y = 1\{partner = B\} \in \{0,1\}$$

on pairs containing A in that subset. We estimated risk ratios using Poisson regression with log link and robust SEs:

$$\log\{\text{E}(Y)\} = \alpha + \beta_{\text{type}}\, 1(\text{IDC or IMC}) + \beta_{\text{sex}}\text{sex} + \beta_{\text{age}}\text{age} + \beta_{\text{year}}\text{year}_{\text{c}} + \sum_{g} \gamma_g 1(ward_{group} = g)$$

The exponentiated sample-type coefficient gives the adjusted RR for the contrast (IDC vs MU or IMC vs MU). *Sex* and *age* were included in all fits, with adjustments for centred calendar year and for a coarsened ward category, treated as fixed effects. Mean-bias-reduced Poisson (brglm2) was used as the primary engine, with standard GLMs as fallback, and non-estimable models after these steps were skipped. For each focal organism A, BH FDR was applied across the set of evaluated partners per contrast, and we report RR, 95% CI, and q (the adjusted p-value). The analysis was performed using the sandwich, (8) lmtest (9), the brglm2 (11) packages in R.

#### 3 Resistance Analysis

The AST data received had already been classified according to the EUCAST (15) guidelines of the time the sample was collected, to be susceptible (S), Intermediate (I) or Resistant (R). Due to significant changes involving the susceptible vs intermediate phenotype over the years in the EUCAST guidelines, only resistance (AST for that antibiotic-bacterial pair = R) was considered as the marker for analysis.

The resistance analysis was performed on the positive samples' dataset, filtered to keep only those samples which had at least one pathogen with measured resistances in them, and keeping only those antibiotics for which the antibiotic-pathogen pair was tested in at least 60% of that bacteria's samples.

The total resistance vs acquired resistance was separated out and analysed separately. Total resistance was defined as the data arising directly from the ASTs obtained. Acquired resistance was defined as (Total Resistance – Inherent resistances), where inherent resistance was defined as the inherent resistance bacteria possess to antibiotics which they were nevertheless tested against during AST calculation. This inherent resistance list was defined based on the concordance of 2 criteria:

1. resistance of >90% of that specific species-antibiotic pair in our dataset, and
2. EUCAST Expected Resistant Phenotypes (Version 1·2, January 2023), or literature detailing inherent resistance for that bacterial species-antibiotic pair.

The file defining inherent vs total resistance is part of the accessory files (always_resistant.xlsx) that are uploaded on Zenodo ((https://doi.org/10.5281/zenodo.18338805).

##### 3.1 Treating Measurements of Antibiotic Resistance as Bernoulli Random Variables

We consider measurements of sample antibiotic resistance to be realizations of a random variable drawn from a Bernoulli probability distribution with parameter $\mathbb{P}(\text{Resistance} = 1)$. Namely, a measurement of resistance is taken to be equivalent to flipping a coin with probability $p$ of producing heads. From $N$ measurements of resistance, we estimate the Bernoulli parameter as

$$\hat{p}_X = \frac{1}{N_X}\sum_{i=1}^{N_X} x_i.$$

Where the $x_i$ are realizations of Bernoulli random variables $X$ which determines resistance and $N_X$ is the number of realizations of $X$. We use the sample variance of the Bernoulli parameter estimator,

$$\text{var}[\hat{p}_X] = \frac{1}{N_X}\hat{p}_X(1 - \hat{p}_X),$$

to construct the standard error of the estimator: $\text{SE}[\hat{p}_X] = \sqrt{\text{var}[\hat{p}_X]}$.

To highlight the differences in antibiotic resistance across samples we also consider $X$ to be a conditional random variable $X = \text{Resistance} \mid Y$. Here $Y$ is taken to be a discrete random variable such as sample type, year of collection, month of collection, patient gender, patient age, medical source of sample, microorganism(s) detected, and microbial concentration. Conditioning in this manner provides a set of Bernoulli parameter estimators over the support of $Y$. For example, if $Y$ is the patient age then the Bernoulli estimator for $X = \text{Resistance} \mid \text{Age}$ becomes a function of the patient age. In principle, we may also condition a set of sample metadata variables: $X = \text{Resistance} \mid Y_1, \ldots, Y_k$. However, the rarity of certain combinations of sample metadata can lead to missing information, large errors, or uniform resistance/susceptibility in a low data regime. For this reason, we take care when conditioned on microorganism(s) detected and other sample metadata to only use samples corresponding to the most frequently detected microorganism(s) in the overall dataset.

##### 3.2 $\chi^2$ Permutation Analysis for Statistical Significance of Resistance and Susceptibility

We define a $\chi^2$ statistic

$$\chi_0^2 = \frac{N_X(\hat{p}_{R|X} - \hat{p}_R)^2}{\hat{p}_R} + \frac{N_X(\hat{p}_{R|X} - \hat{p}_R)^2}{1 - \hat{p}_R},$$

where the null hypothesis is equivalency in the number of measured resistant and susceptible samples with metadata $X$ vs. the number expected if resistance and susceptibility were uniformly distributed across all sample metadata. We permutate all results of resistance testing by sampling permutations with replacement. We denote each such permutation of the samples by $\pi \in \Pi$, respectively. For each permutation we calculate Bernoulli parameter estimators which we denote by $\hat{p}_{\pi[R]|X}$. The resulting $\chi^2$ statistics are

$$\chi^2 = \frac{N_X(\hat{p}_{\pi[R]|X} - \hat{p}_R)^2}{\hat{p}_R} + \frac{N_X(\hat{p}_{\pi[R]|X} - \hat{p}_R)^2}{1 - \hat{p}_R}.$$

Consequently, we define the *p*-value to be

$$p = \frac{1}{|\Pi|} \sum_{\pi \in \Pi} \mathbb{1}\{\chi^2(\pi) \geq \chi_0^2\},$$

which is the fraction of occurrences in the sampled permutations that the $\chi^2$ statistic is greater than the unpermuted $\chi^2$ statistic.

##### 3.3 Calculating Mutual Information

Mutual information (MI) quantifies how much a metadata variable reduces uncertainty about resistance. MI is reported in bits, where 0 indicates no predictive value and 1 bit represents the maximum uncertainty reduction possible for a binary outcome. Because resistance prevalence is typically imbalanced (that is, not 50-50), the theoretical upper bound is well below 1 bit; thus, MI values in the range of 0.2–0.3 bits already reflect meaningful predictive information.

Mutual information is directly computed from the Bernoulli parameter estimators:

$$I(R, X) = \sum_{x \in X} \left\{ (1 - \hat{p}_R) \log_2 \left[ \frac{\mathbb{P}_{R,X}(0, x)}{(1 - \hat{p}_R)\mathbb{P}_X(x)} \right] + \hat{p}_R \log_2 \left[ \frac{\mathbb{P}_{R,X}(1, x)}{\hat{p}_R \mathbb{P}_X(x)} \right] \right\}$$

where $\hat{p}_R$ is the Bernoulli parameter estimator of the unconditioned sample resistance and $\mathbb{P}_{R,X}(\cdot, x)$ is the joint probability distribution of resistance and a particular sample metadata variable $X$.

We calculate a measure of variance by treating mutual information as an expectation value of a random variable:

$$I(R, X) = \mathbb{E} \log_2 \left[ \frac{\mathbb{P}_{R,X}}{\mathbb{P}_R \mathbb{P}_X} \right],$$

which in turn provides that the variance is

$$\mathrm{var}[I] = \mathbb{E} \log_2^2 \left[ \frac{\mathbb{P}_{R,X}}{\mathbb{P}_R \mathbb{P}_X} \right] - \left( \mathbb{E} \log_2 \left[ \frac{\mathbb{P}_{R,X}}{\mathbb{P}_R \mathbb{P}_X} \right] \right)^2.$$

Hence, we associate to each value of mutual information a measure of error in the form of the standard deviation of the associated random variable.

##### 3.4 Error Propagation via Delta Method for Log-Fold Change

We define a random variable $Z = \log_2(\hat{p}_X/\hat{p}_Y)$ which is the log-fold change between two Bernoulli parameter estimators $\hat{p}_X$ and $\hat{p}_Y$. Assuming that $\hat{p}_X$ and $\hat{p}_Y$ are independent, the sample variance of $Z$ can be approximated by the delta method as

$$\begin{aligned}\mathrm{var}[Z] &\approx \left(\frac{\partial Z}{\partial \hat{p}_X}\right)^2 \mathrm{var}[\hat{p}_X] + \left(\frac{\partial Z}{\partial \hat{p}_Y}\right)^2 \mathrm{var}[\hat{p}_Y] \\ &= \frac{1}{\log^2 2}\left[\left(\frac{\partial \log \hat{p}_X}{\partial \hat{p}_X}\right)^2 \mathrm{var}[\hat{p}_X] + \left(\frac{\partial \log \hat{p}_Y}{\partial \hat{p}_Y}\right)^2 \mathrm{var}[\hat{p}_Y]\right] \\ &= \frac{1}{\log^2 2}\left(\frac{\mathrm{var}[\hat{p}_X]}{\hat{p}_X^2} + \frac{\mathrm{var}[\hat{p}_Y]}{\hat{p}_Y^2}\right).\end{aligned}$$

Consequently, the standard error of $Z$ is taken to be

$$\mathrm{SE}[Z] \approx \frac{1}{\log 2}\sqrt{\frac{\mathrm{var}[\hat{p}_X]}{\hat{p}_X^2} + \frac{\mathrm{var}[\hat{p}_Y]}{\hat{p}_Y^2}},$$

where the sample variance of the Bernoulli parameter estimator is as previously defined.

##### 3.5 $\chi^2$ Permutation Analysis for Statistical Significance of Bernoulli Estimator Differences

We define a $\chi^2$ statistic

$$\chi_0^2 = \frac{(\hat{p}_X - \hat{p}_Y)^2}{\hat{p}_Y}$$

where the null hypothesis is that the Bernoulli parameter estimators are equivalent. We permutate all results of resistance testing for bimicrobial and monomicrobial samples by sampling permutations with replacement. We denote each such permutation of the bimicrobial and monomicrobial samples by $(\pi_1, \pi_2) \in \Pi$, respectively. For each permutation we calculate Bernoulli parameter estimators which we denote by $\hat{p}_{\pi_1[X]}$ and $\hat{p}_{\pi_2[Y]}$. The resulting $\chi^2$ statistics are

$$\chi^2(\pi_1, \pi_2) = \frac{(\hat{p}_{\pi_1[X]} - \hat{p}_{\pi_2[Y]})^2}{\hat{p}_{\pi_2[Y]}}.$$

Consequently, we define the *p*-value to be

$$p = \frac{1}{|\Pi|}\sum_{(\pi_1,\pi_2)\in\Pi} \mathbb{1}\{\chi^2(\pi_1, \pi_2) \geq \chi_0^2\},$$

which is the fraction of occurrences in the sampled permutations that the $\chi^2$ statistic is greater than the unpermuted $\chi^2$ statistic.

#### Supplementary Figures

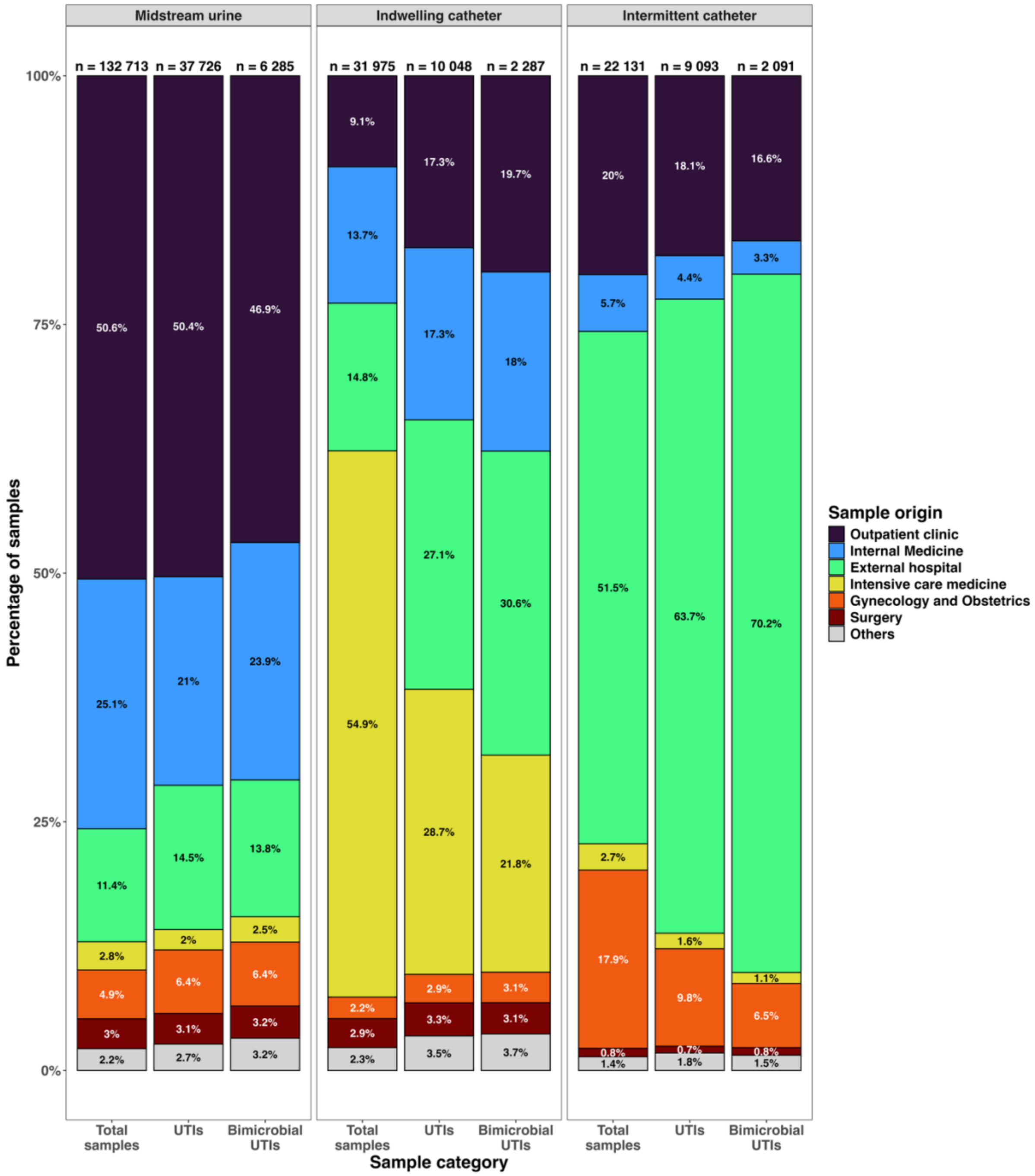


**Figure S1 – Origin of urine samples, separated by sample type and category of samples.** This figure shows the different origins of the samples assessed in this study, comparing the sample types (midstream urine, indwelling catheters, intermittent catheters), for total samples, samples classified as UTIs, and bimicrobial UTIs. See ward data files for complete lists.

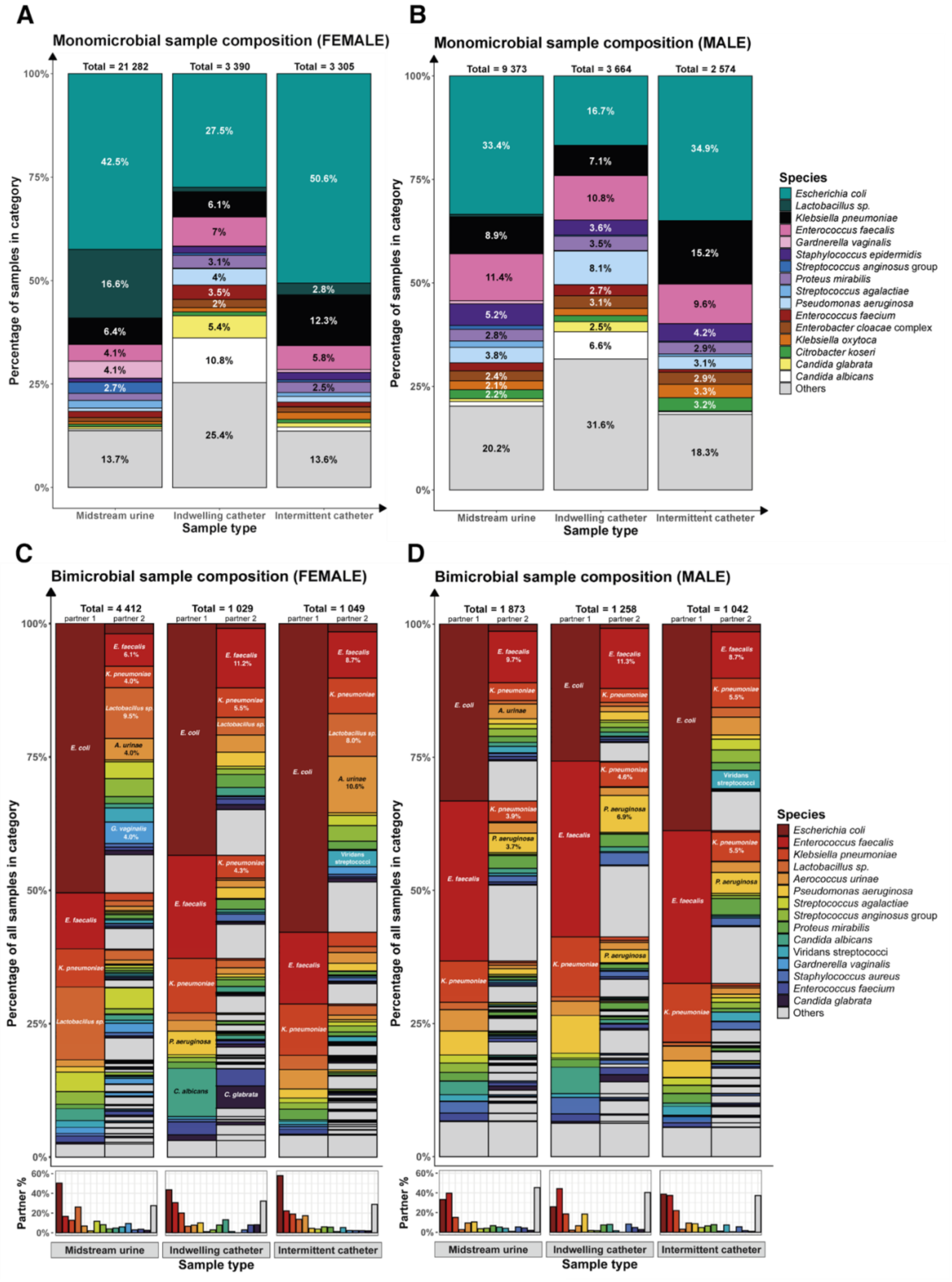


**Figure S2 – Composition of monomicrobial and bimicrobial UTI samples, based on urine sample type, across male and female patients.** This figure compares the microbial composition of monomicrobial UTI samples in A) female patients and B) male patients, followed by the microbial composition of bimicrobial UTI samples in C) female patients and D) male patients. In A) and B), microbes which make up >2% share of the total have their share labelled per sample type. In C) and D), the top 5 microbial pairs are labelled, and the top 3 have their percentage share mentioned. The inset figures at the bottom refer to the percentage of samples, per sample type, that have the organisms graphed as a partner. See mono_combo and merged_combination data files for complete lists.

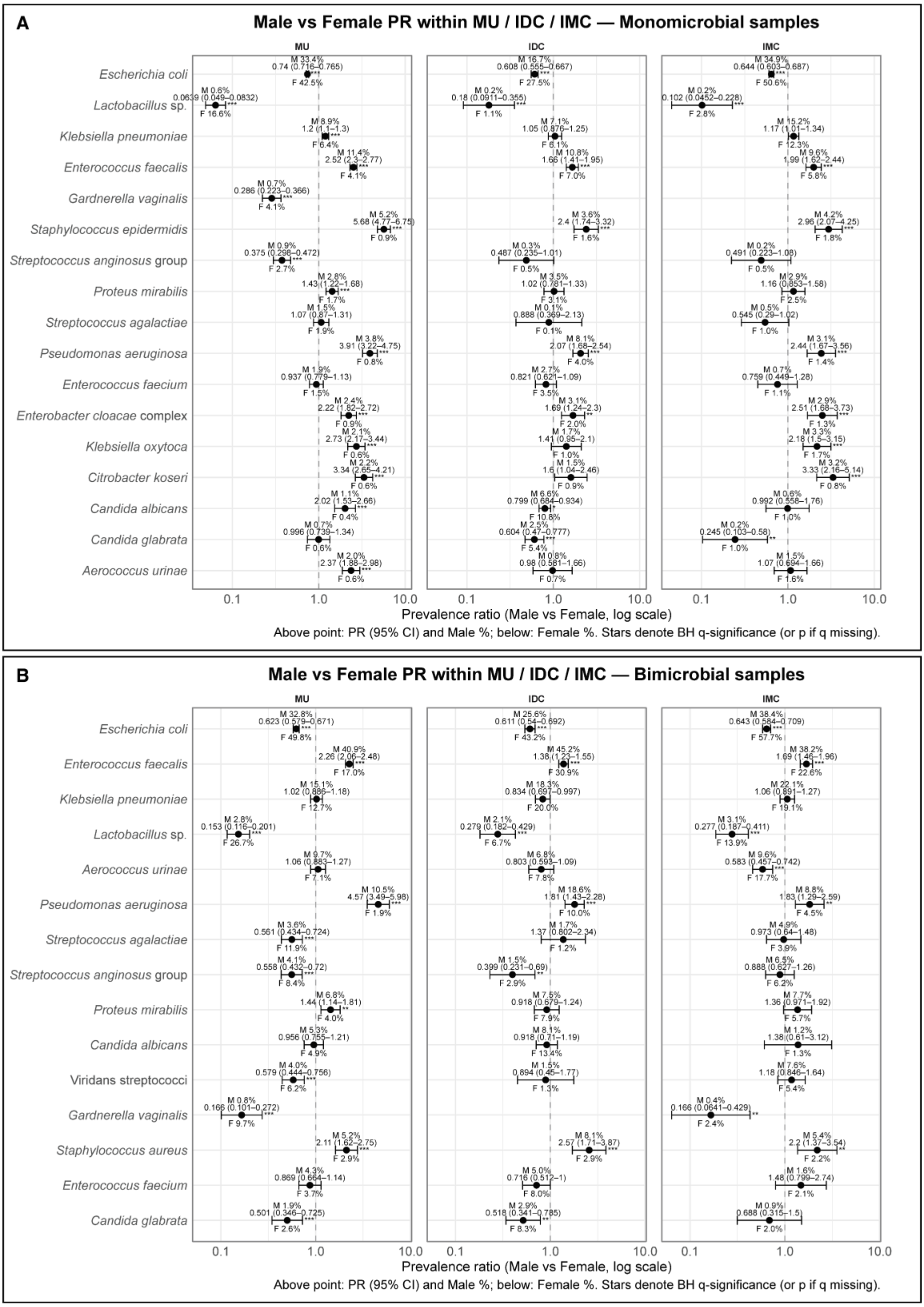


**Figure S3 – Male vs Female PRs within sample types**. This figure shows the Male vs Female PRs (robust log-Poisson) with 95% CIs for each sample type, for selected pathogens (like Figure 2) for A) Monomicrobial samples, and B) Bimicrobial samples. Labels show **%** positive in males and females within each type, along with the PRs [CI]. Significance has been estimated using BH-adjusted q-values ($q$<0·05 *, <0·01 **, <0·001 ***). See data file Prevalence_Ratio_Data.xlsx for complete lists.

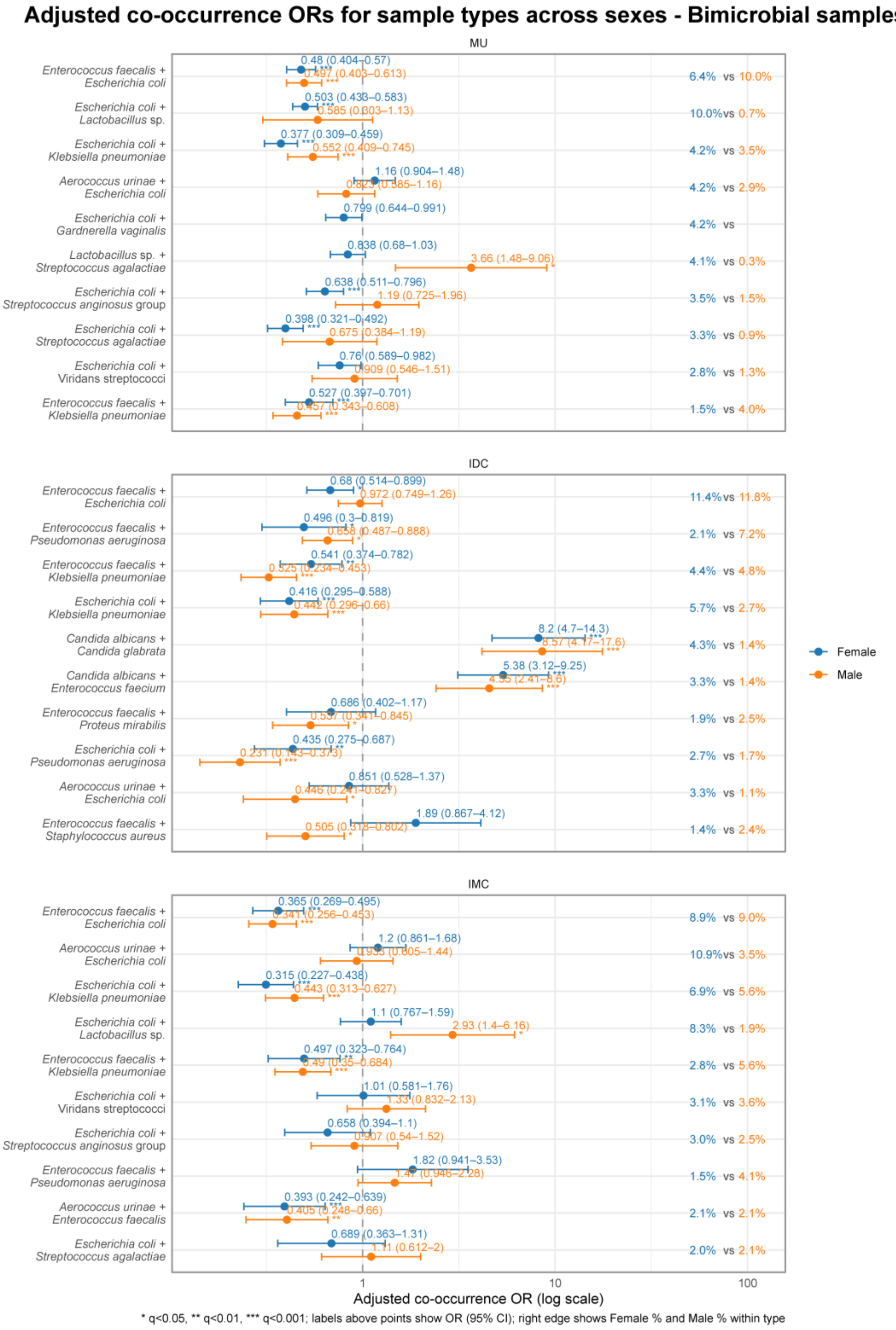


**Figure S4 – Adjusted co-occurrence odds ratios ($OR_{sym}$) by sample type (bimicrobial UTIs, stratified by sex).** Each panel shows $OR_{sym}$ (point) with 95% CI (horizontal line) on a log scale for organism pairs within each sample type, which was stratified by sex. CIs use the mean SE on the log scale, and $q$ is the minimum BH-adjusted $p$ across directions. Right-edge labels show the pair prevalence (%) (co-appearances / total samples) per sex – female data points are coloured blue, while male data points are coloured orange. Models are adjusted for age, ward groups and year of collection Asterisks mark significance ($q$<0·05 *, <0·01 **, <0·001 ***).

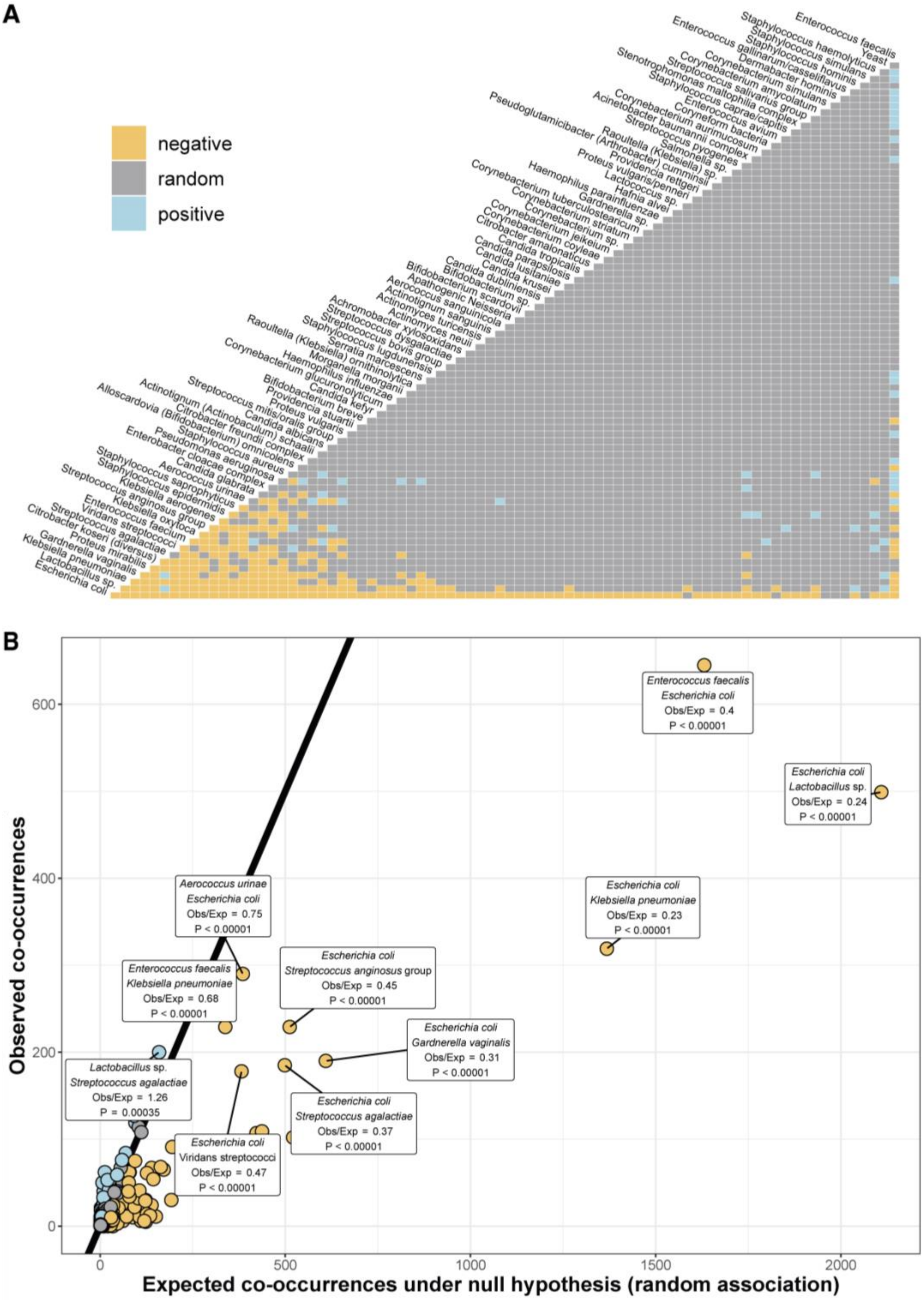


**Figure S5 – Classical co-occurrences in UTIs from midstream urine samples agree with all-sample OR values.** This figure compares the observed co-occurrence of microbes with their expected co-occurrences, across <u>all infection sites</u> where positive interactions are coloured blue, negative interactions are coloured gold, and random interactions are coloured grey. A) This figure qualitatively summarizes the associations of all pairs in the dataset, via the species-co-occurrence matrix, while in B), the same data is represented as an observed vs expected co-occurrences, with the highest observed pairs (>160) labelled with their details – the microbial species, the observed to expected ratios, and the significance of interaction, which can be used as P values. Co-occurrences for the other sample types (IDC, IMC) and the overall data are present in data file Classical_cooccur.xlsx.

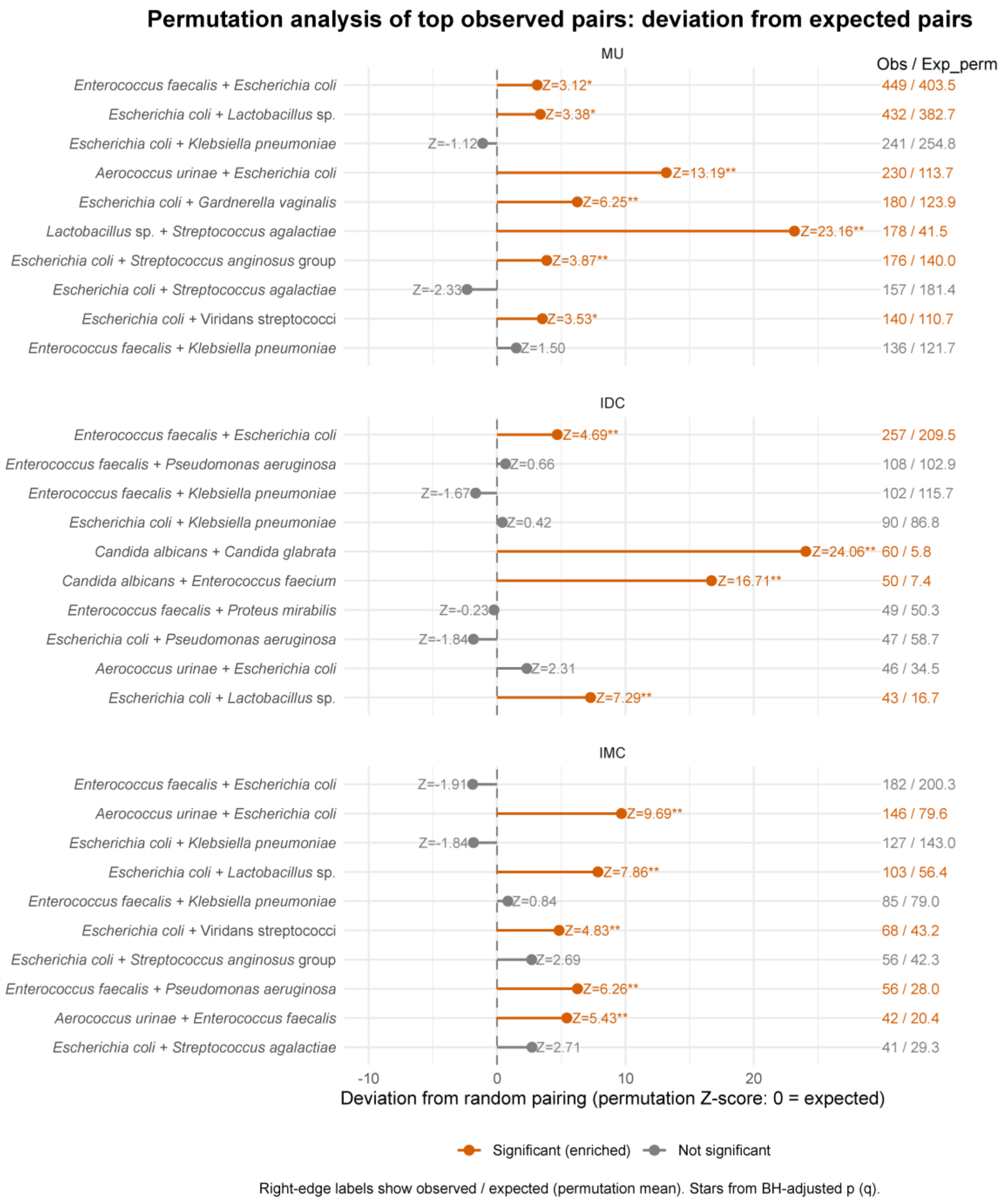


**Figure S6 – Degree-preserving permutation results in bimicrobial samples.** For each sample type, "lollipop plots" show the Top 10 pairs by prevalence in that stratum. The x-axis is the permutation Z-score, the points mark the Z value and the significance. Right-edge labels show the observed/expected values. All significantly enhanced rows (implying that actual prevalence of a pair is more than that could be explained by prevalence alone) are coloured orange, and the significantly depleted ones are coloured blue (there are none in the shown subset). q values are BH-adjusted within material. Asterisks mark significance (*q*<0·05 *, <0·01 **, <0·001 ***). Raw data is present in the data file Permutations_Pair_enrichment.xlsx.

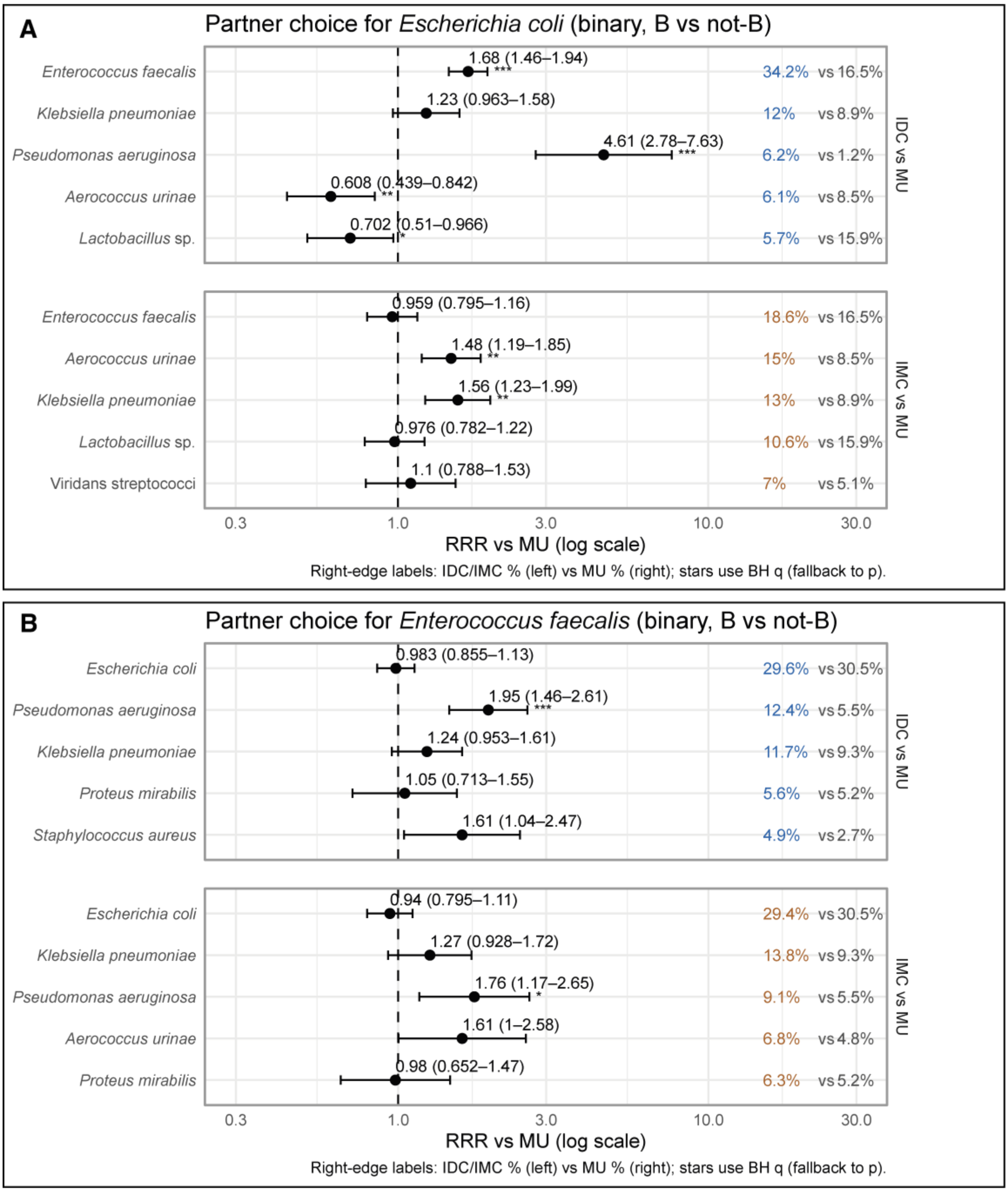


**Figure S7 – Partner choice analysis for *E. coli and E. faecalis* as partners in bi-microbial UTIs (binary "B vs not-B").** Points show adjusted risk ratios (RR) and 95% CIs on a log scale for IDC vs MU and IMC vs MU, estimated among bimicrobial pairs that include A) *E. coli* and B) *E. faecalis*. Within each contrast, the top 5 partners were selected by frequency. Asterisks mark BH-adjusted significance ($q$<0·05 *, <0·01 **, <0·001 ***). Right-edge labels report sample type% (coloured; IDC or IMC) versus MU % (grey) for "partner = B" among the respective pairs.

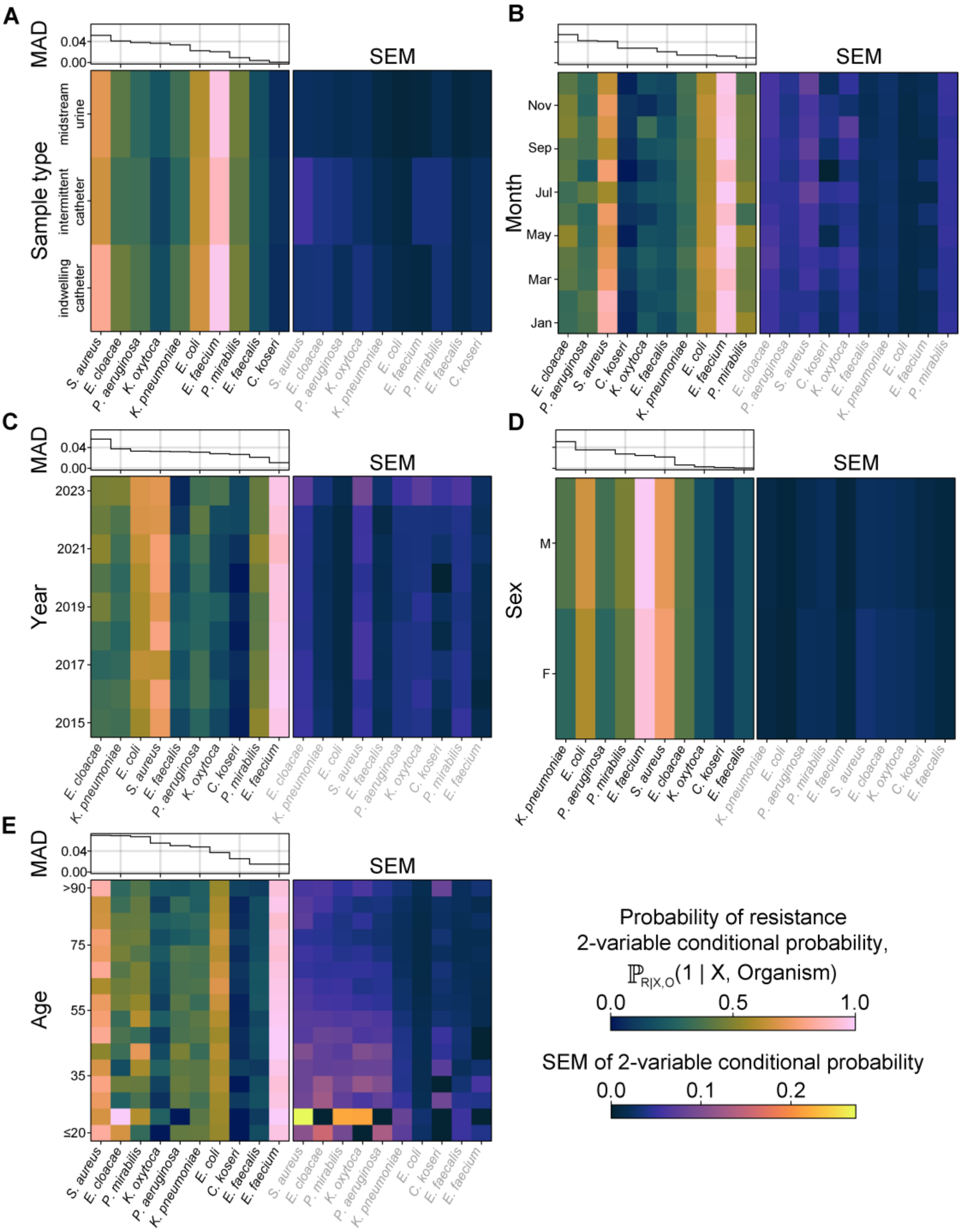


**Figure S8 – Antibiotic resistance prevalence for organisms varies across sample parameters of patient characteristics, sample type and sampling time.** Combined probabilistic analysis of resistance across organisms and other sample parameters was performed. Resistance of the 10 most abundant organisms within monomicrobial samples conditioned on 2 of 8 sample metadata variables is treated as a Bernoulli random variable with parameters shown in (A)–(E) (left) for monomicrobial samples with acquired resistances. Corresponding standard errors of the mean are shown in (A)–(E) (right). Organisms are ordered by the descending median absolute deviation (MAD) shown in (A)–(E) top. Raw data is present in the data file Resistance_Tables.xlsx.

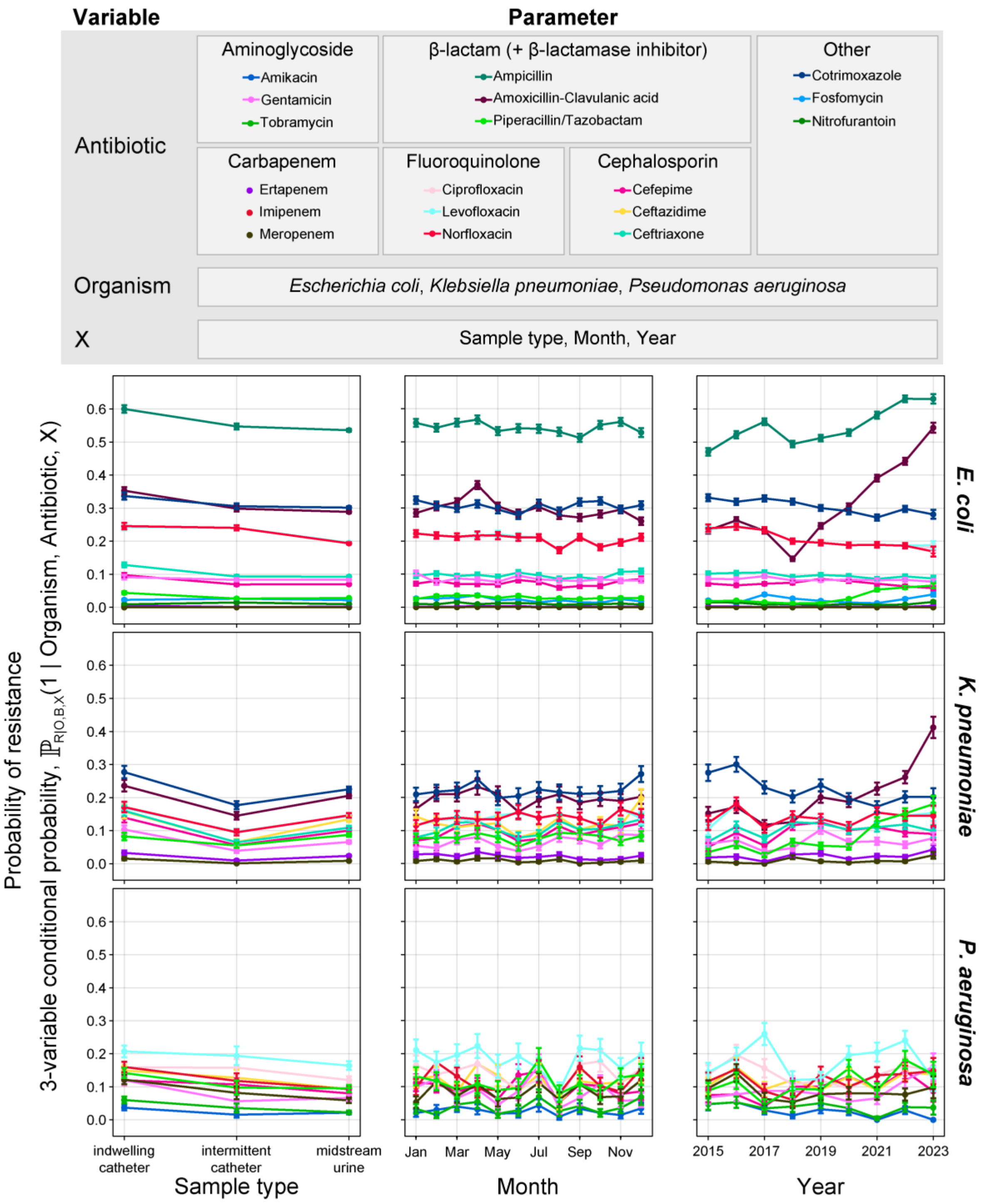


**Figure S9 – Antibiotic resistance against commonly used antibiotics changes over time, across sample types and for different organisms.** Resistance conditioned on antibiotic tested and sample material (left), month of sample collection (centre), or year of sample collection (right), is treated as a Bernoulli random variable with parameters as shown for *Escherichia coli* (top), *Klebsiella pneumoniae* (middle), and *Pseudomonas aeruginosa* (bottom). All error bars shown are standard error of the mean (SEM). The antibiotics shown in this analysis are those that were part of the ASTs for >60% of all the ASTs for that organism consistently. Raw data of each subpanel is present in the data file Resistance_Tables.xlsx.

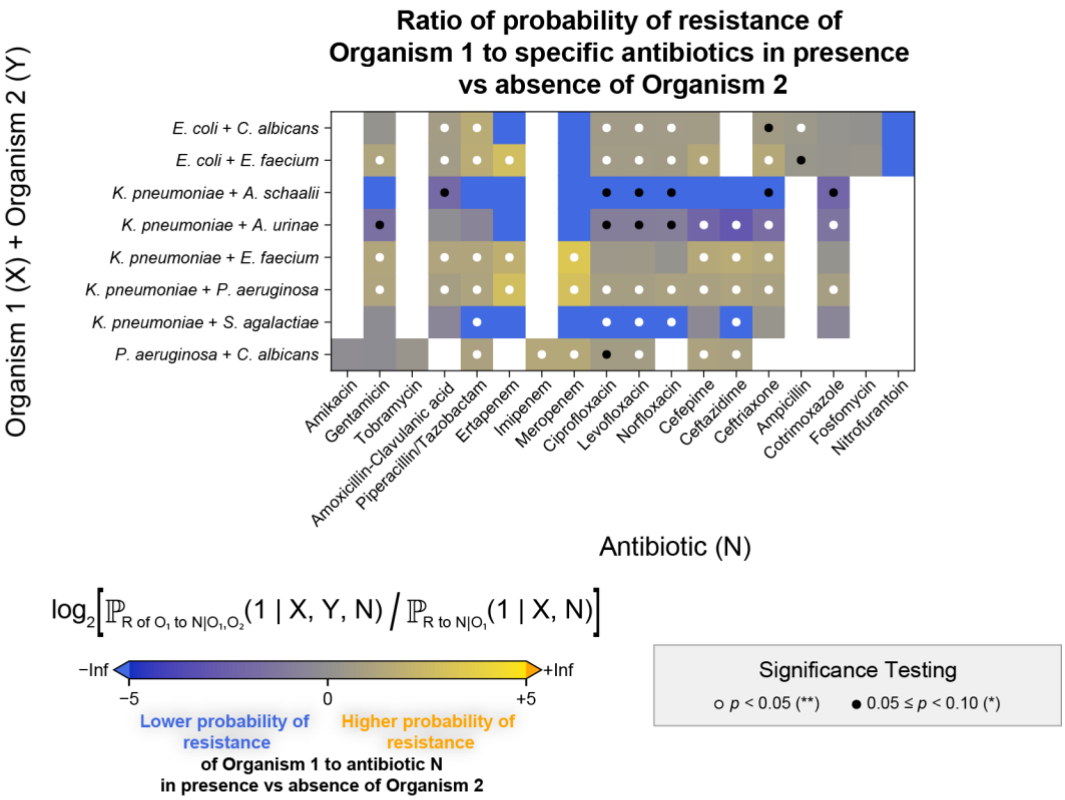


**Figure S10** – **Antibiotic resistance changes in the presence of partners compared to their absence for pathogen combinations with significant differences.**
The log fold change in resistance for *E. coli* in the presence vs absence of *C. albicans* and *E. faecium*, for *K. pneumoniae* in the presence vs absence of *A. schaalii*, *A. urinae*, *E. faecium*, *P. aeruginosa* and *S. agalactiae*, and for *P. aeruginosa* in the presence vs absence of *C. albicans* has been graphed above, with the impact on individual resistances substantiated. Significance testing was performed using a double permutation test against a zero-change null hypothesis as described in the Methods. Raw data is present in data file Resistance_Tables.xlsx

#### Supplementary Tables

**Table S1: Sex of the sample provider, and number of species in each sample, separated by sample type.**

| Sample Type | Sex | All UTI n | 1 species n (%) | 2 species n (%) | >=3 species n (%) |
|---|---|---|---|---|---|
| **MU** | Male | 11554 | 9373 (81·1%) | 1873 (16·2%) | 308 (2·7%) |
| **MU** | Female | 26172 | 21282 (81·3%) | 4412 (16·9%) | 478 (1·8%) |
| **IDC** | Male | 5379 | 3664 (68·1%) | 1258 (23·4%) | 457 (8·5%) |
| **IDC** | Female | 4669 | 3390 (72·6%) | 1029 (22·0%) | 250 (5·4%) |
| **IMC** | Male | 4266 | 2574 (60·3%) | 1042 (24·4%) | 650 (15·2%) |
| **IMC** | Female | 4827 | 3305 (68·5%) | 1049 (21·7%) | 473 (9·8%) |

**Table S2: Prevalence Ratios for polymicrobial nature of samples – adjusted for age, sex, ward and year of collection**

| Samples | Contrast | PR [95% CI] | P values |
|---|---|---|---|
| **All positive samples (Bacteriuria + UTI)** | IDC vs MU | 1.54 [1.48–1.60] | 4.47E-100 |
| | IMC vs MU | 1.53 [1.47–1.58] | 3.52E-119 |
| **UTI Samples (Total CFU>= $10^5$)** | IDC vs MU | 1.63 [1.56–1.70] | 1.17E-115 |
| | IMC vs MU | 1.62 [1.56–1.69] | 2.36E-128 |

**Table S3: RDA analysis data – marginal and pairwise, showing the differences between the sample types in terms of microbial composition.**

| **Marginal RDA Tests** | | | | |
|---|---|---|---|---|
| **Term** | **Degrees of freedom** | **F** | **Pr(>F)** | **Partial adj $R^2$** |
| **Sample type** | 2 | 194.71 | 0.001 | 0.0068 |
| **Sex** | 1 | 491.56 | 0.001 | 0.00861 |
| **Age** | 1 | 304.22 | 0.001 | 0.00532 |
| **Year** | 1 | 40.42 | 1 | 6.92E-04 |
| **Ward Groups** | 5 | 59.52 | 0.001 | 0.00513 |
| **Overall** | NA | NA | NA | 0.03554 |

| Levels | N | S | F | $R^2$adj | p (BH adjusted) |
|---|---|---|---|---|---|
| **MU vs IDC** | 46039 | 27 | 331.43662 | 0.006902 | 0·001 |
| **MU vs IMC** | 45262 | 27 | 53.4511094 | 0.00112405 | 0·001 |
| **IMC vs IDC** | 18442 | 28 | 187.24097 | 0.00977513 | 0·001 |

**Table S4: This table shows the ORs of species pairs which are significant but non-concordant between the bimicrobial cohort and ORs from the entire UTI database: all flips were from depleted in bimicrobial samples to enriched in polymicrobial samples**

| Non-concordance between bimicrobial ORs and all-sample ORs: 4 pair flips | | | | | | | |
|---|---|---|---|---|---|---|---|
| Sample Type | Pair | Bimicrobial | | | All samples | | |
| | | OR [95% CI] | $q_{min}$ | Sample% | OR [95% CI] | $q_{min}$ | Sample% |
| **IDC** | *Actinotignum schaalii* \| *Enterococcus faecalis* | 0·325 [0·125-0·846] | 4·08E-02 | 0·2% | 3·242 [2·041-5·150] | 3·50E-06 | 0·4% |
| **IDC** | *Aerococcus urinae* \| *Enterococcus faecalis* | 0·374 [0·250-0·559] | 7·89E-06 | 1·5% | 1·648 [1·292-2·101] | 1·55E-04 | 1·2% |
| **IMC** | *Aerococcus urinae* \| *Enterococcus faecalis* | 0·399 [0·282-0·564] | 9·53E-07 | 2·1% | 2·319 [1·958-2·746] | 3·85E-21 | 2·8% |
| **IMC** | *Aerococcus urinae* \| *Streptococcus anginosus* group | 0·250 [0·101-0·621] | 7·95E-03 | 0·2% | 1·659 [1·221-2·254] | 4·89E-03 | 0·6% |

**Table S5: This table shows the species pairs for which the degree preserving permutations are significant but non-concordant to significant adjusted ORs: all flips from depleted ORs to enriched permutations**

| Non-concordance between bimicrobial ORs and degree-preserving permutations | | | | | | | | |
|---|---|---|---|---|---|---|---|---|
| **Sample Type** | **Pair** | **Num** | **Sample %** | **Expected perm** | **Z perm** | **q perm** | **OR [95%CI]** | **q OR** |
| **IDC** | *Aerococcus urinae* \|*Enterococcus faecalis* | 33 | 1·5% | 18·1 | 3·92 | 5·29E-03 | 0·374 [0·250-0·560] | 7·89E-06 |
| **IDC** | *Enterobacter cloacae* complex \|*Enterococcus faecalis* | 33 | 1·5% | 15·7 | 4·93 | 3·08E-03 | 0·566 [0·370-0·866] | 2·03E-02 |
| **IMC** | *Aerococcus urinae* \|*Enterococcus faecalis* | 42 | 1·9% | 20·4 | 5·43 | 2·78E-03 | 0·399 [0·282-0·564] | 9·53E-07 |
| **MU** | *Aerococcus urinae*\| *Enterococcus faecalis* | 70 | 1.2% | 30.2 | 7.91 | 2.29E-03 | 0.383 [0.290–0.507] | 1.74E-10 |
| **MU** | *Candida albicans*\| *Enterococcus faecalis* | 29 | 0.5% | 16.8 | 3.17 | 2.86E-02 | 0.272 [0.183–0.406] | 1.02E-09 |
| **MU** | *Enterococcus faecalis*\| *Escherichia coli* | 449 | 7.5% | 403.5 | 3.12 | 1.63E-02 | 0.491 [0.430–0.561] | 5.55E-24 |
| **MU** | *Escherichia coli*\| *Gardnerella vaginalis* | 180 | 3.0% | 123.9 | 6.25 | 2.29E-03 | 0.765 [0.619–0.944] | 2.36E-02 |
| **MU** | *Escherichia coli*\| *Lactobacillus sp.* | 432 | 7.2% | 382.7 | 3.38 | 1.06E-02 | 0.501 [0.433–0.578] | 6.39E-20 |
| **MU** | *Escherichia coli*\| *Streptococcus anginosus* group | 176 | 2.9% | 140.0 | 3.87 | 2.29E-03 | 0.719 [0.584–0.884] | 4.28E-03 |
| **MU** | *Gardnerella vaginalis*\| *Lactobacillus* sp. | 79 | 1.3% | 43.3 | 5.91 | 2.29E-03 | 0.284 [0.212–0.380] | 3.23E-16 |
| **MU** | *Gardnerella vaginalis*\| *Streptococcus agalactiae* | 46 | 0.8% | 19.0 | 6.55 | 2.29E-03 | 0.479 [0.340–0.675] | 8.75E-05 |
| **MU** | *Gardnerella vaginalis*\| *Streptococcus anginosus group* | 28 | 0.5% | 14.3 | 3.81 | 3.98E-03 | 0.636 [0.419–0.963] | 3.85E-02 |
| **MU** | *Klebsiella pneumoniae*\| *Proteus mirabilis* | 26 | 0.4% | 15.1 | 2.96 | 4.50E-02 | 0.537 [0.353–0.815] | 6.97E-03 |
| **MU** | *Klebsiella pneumoniae*\| *Pseudomonas aeruginosa* | 29 | 0.5% | 13.4 | 4.43 | 5.49E-03 | 0.650 [0.434–0.974] | 4.27E-02 |
| **MU** | *Lactobacillus* sp.\|*Streptococcus anginosus group* | 44 | 0.7% | 29.4 | 2.88 | 4.71E-02 | 0.291 [0.210–0.404] | 8.05E-13 |
| **MU** | *Streptococcus agalactiae*\| *Streptococcus anginosus* group | 27 | 0.4% | 4.2 | 11.49 | 2.29E-03 | 0.508 [0.331–0.779] | 3.65E-03 |
| **MU** | *Streptococcus agalactiae*\| Viridans streptococci | 17 | 0.3% | 2.8 | 8.71 | 2.29E-03 | 0.473 [0.283–0.790] | 6.46E-03 |

#### References:


1. R Core Team. R: A Language and Environment for Statistical Computing [Internet]. Vienna, Austria: R Foundation for Statistical Computing; 2023. Available from: https://www.R-project.org/
2. Wickham H, Averick M, Bryan J, Chang W, McGowan LD, François R, et al. Welcome to the Tidyverse. J Open Source Softw. 2019 Nov 21;4(43):1686.
3. Slowikowski K. ggrepel: Automatically Position Non-Overlapping Text Labels with “ggplot2.” 2024.
4. Wilke CO. cowplot: Streamlined Plot Theme and Plot Annotations for ‘ggplot2.’ J Open Source Softw. 2019;4(35):1071.
5. Garnier S, Ross N, Rudis R, Camargo A, Sciaini M, Scherer C. viridis - Colorblind-Friendly Color Maps for R. J Open Source Softw. 2021;6(64):2973.
6. Neuwirth E. RColorBrewer: ColorBrewer Palettes. R Package Version 11-2 [Internet]. 2014; Available from: https://CRAN.R-project.org/package=RColorBrewer
7. Krzywinski M. Designing for Color Blindness—15-Colour Palette [Internet]. 2025. Available from: https://mk.bcgsc.ca/colorblind/palettes.mhtml
8. Zeileis A, Köll S, Graham N. Various Versatile Variances: An Object-Oriented Implementation of Clustered Covariances in R. J Stat Softw. 2020 Oct 7;95:1–36.
9. Zeileis A, Hothorn T. Diagnostic Checking in Regression Relationships. R News. 2002;2(3):7–10.
10. Oksanen J, Simpson GL, Blanchet FG, Kindt R, Legendre P, Minchin PR, et al. vegan: Community Ecology Package [Internet]. 2024. Available from: https://CRAN.R-project.org/package=vegan
11. Kosmidis I. brglm2: Bias Reduction in Generalized Linear Models [Internet]. 2025. Available from: https://CRAN.R-project.org/package=brglm2
12. Griffith DM, Veech JA, Marsh CJ. cooccur: Probabilistic Species Co-Occurrence Analysis in R. J Stat Softw. 2016 Feb 9;69:1–17.
13. Barrett T, Dowle M, Srinivasan A, Gorecki J, Chirico M, Hocking T. data.table: Extension of `data.frame` [Internet]. 2024. Available from: https://CRAN.R-project.org/package=data.table
14. Bengtsson H. A Unifying Framework for Parallel and Distributed Processing in R using Futures. R J. 2021;13(2):208–27.
15. eucast: EUCAST [Internet]. [cited 2025 Sept 2]. Available from: https://www.eucast.org/
